# Does the Heart Fall Asleep? – Diurnal Variations of Heart Rate Variability in Patients with Disorders of Consciousness

**DOI:** 10.1101/2022.02.07.22270241

**Authors:** Monika Angerer, Frank H. Wilhelm, Michael Liedlgruber, Gerald Pichler, Birgit Angerer, Monika Scarpatetti, Christine Blume, Manuel Schabus

**Author notes:** Correspondence (M.A.), (Ma.S.). Christine Blume and Manuel Schabus contributed equally to this work. (F.W.); (M.L.). (G.P.); (Mo.S.).

## Abstract

The current study investigated heart rate (HR) and heart rate variability (HRV) across day and night in patients with disorders of consciousness (DOC). We recorded 24-h ECG in 26 patients with DOC (i.e., unresponsive wakefulness syndrome [UWS; *n*=16] and (exit) minimally conscious state [(E)MCS; *n*=10]). To examine diurnal variations, HR and HRV indices in the time, frequency, and entropy domains were computed for periods of clear day- (forenoon: 8am-2pm; afternoon: 2pm-8pm) and nighttime (11pm-5am). Results indicate that patients’ interbeat intervals (IBIs) were larger during the night than during the day indicating HR slowing. Additionally, higher HRV entropy was associated with higher EEG entropy during the night. Patients in UWS showed larger IBIs compared to patients in (E)MCS, and patients with non-traumatic brain injury showed lower ECG entropy than patients with traumatic brain injury. Thus, cardiac activity varies with a diurnal pattern in patients with DOC and can differentiate between patients’ diagnoses and etiologies. Moreover, also the interaction of heart and brain appears to follow a diurnal rhythm. Thus, HR and HRV seem to mirror the integrity of brain functioning and consequently might serve as supplementary measures for improving the validity of assessments in patients with DOC.

## 1. Introduction

Severe brain injury can cause coma and, upon recovery, changes in consciousness often persist. These states are subsumed under the term ‘disorders of consciousness’ (DOC). In a simplified approach, two major components are thought to be necessary for consciousness: wakefulness (i.e., the level of arousal) and awareness of the environment and the self (i.e., contents of consciousness) [1]. In patients living with DOC, wakefulness is preserved but awareness is only intermittently present or completely absent. More specifically, while patients with an unresponsive wakefulness syndrome (UWS) show some return of arousal (i.e., phases of sleep [closed eyes] and wakefulness [open eyes]) without signs of awareness during behavioral assessment, patients in a minimally conscious state (MCS) show inconsistent but reproducible signs of awareness that can be differentiated from reflexive behavior (e.g., response to commands, visual pursuit, intentional communication) [2,3]. If patients can communicate functionally and use objects adequately, their state is denoted as emergence from minimally conscious state (EMCS) [4]. Thus, while patients with UWS are assumed to be unconscious, patients in MCS and EMCS are assumed to be (minimally) conscious.

Clinical diagnoses are usually based on observations of the patients’ behavior using for instance the Glasgow Coma Scale [5] for acute situations or the Coma Recovery Scale-Revised (CRS-R) [6] for tracing their development during recovery. Unfortunately, behavioral assessments involve the risk of underestimating the level of consciousness. This is because patients may be unable to respond behaviorally (and thus give evidence of their consciousness) for example due to sensory or motor impairments. Finally, the fluctuating levels of consciousness carry the risk of examinations taking place during windows of unconsciousness [7]. Consequently, the absence of evidence for consciousness must not be mistaken for evidence of its absence. Hence, distinguishing between UWS and (E)MCS continues to be a challenge in clinical practice, and the rate of misdiagnoses is high (i.e., ∼40%) [8]. Approaches based on neuroimaging methods such as functional magnetic resonance imaging (fMRI) or electroencephalography (EEG) have been used as additional tools to improve the validity of DOC diagnoses [9,10]. However, they require special expertise, come with high time and financial requirements and often rely on tasks that are still challenging for patients [11]. Therefore, researchers and clinicians are looking for alternative or adjunct measures that (i) do not rely on patients’ behavioral responses, (ii) are time and cost efficient, (iii) and can be easily applied at beside. Last (iv), they should be useable during longer-term recordings thereby taking the problem of fluctuating consciousness levels over the circadian day into account.

Among these measures, heart rate (HR) and heart rate variability (HRV) have been suggested to fulfill these criteria [12]. Specifically, HR indicates the average time interval between adjacent heartbeats (i.e., interbeat interval; IBI), while HRV quantifies the variability in these time intervals. These variations occur at different frequencies and reflect dynamics of autonomic nervous system regulation, being related to sympathetic and parasympathetic activity, breathing, thermo-, and blood pressure regulation, as well as changes in the vasomotor, and renin-angiotensin system [13; cf. *Box 1*]. Furthermore, HR/HRV may mirror the interaction between the (injured) brain and the heart and thus represent a ‘peripheral’ window to ‘central’ functioning. The neural structure that enables the bidirectional communication of the heart and central nervous system has been described as the central autonomic network, which has been suggested to be involved in cognitive, emotional, and autonomic regulation, and linked to HR/HRV and cognitive performance [14].

In patients with severe brain injury, a decrease of HRV parameters in the time and frequency domain has been associated with a worse clinical outcome [e.g., 15,16]. When looking at HRV entropy, it has been shown that patients with UWS show lower approximate entropy (ApEn) values than healthy controls [17]. Furthermore, higher complexity values, which were associated with higher CRS-R scores, were observed in patients in MCS as compared to patients with UWS [12]. Interestingly, no differences between patients with UWS and healthy controls has been found in any of the linear parameters (i.e., root mean square of successive differences between adjacent heartbeats [RMSSD], standard deviation of IBIs [SDRR], ratio between low and high frequencies [LF/HF ratio]) [17].

Importantly, like other physiological signals (such as body temperature and hormone secretion), HR/HRV in healthy populations shows strong alterations between sleep and wakefulness [18-20]; so-called circadian variations (i.e., rhythms with a period length of approximately 24 h). In patients with severe brain injury, it has been shown that circadian temperature, melatonin, or motoric activity rhythms often deviate from the healthy norm, and that a better integrity of the patients’ circadian rhythm is associated with a better clinical state [7,21,22]. Until now, it remains unclear how day vs. night variation affects HR/HRV of patients with DOC and to what degree the existence of diurnal variation is linked to the patients’ clinical state.

Thus, our aim was to explore HRV in patients with DOC in the time, frequency and entropy domains during day and night periods. Specifically, as light is the most important zeitgeber for the internal biological clock [23], we also took the lighting conditions in the patients’ room into account.Additionally, we investigated whether there is an association between the patients’ heart and brain activity.

### Box 1.

Methodologically, HRV can be evaluated in the frequency, time, and entropy domains. In the frequency domain, researchers usually compute a fast Fourier transform for three specific frequency bands that can be related to functions of the autonomic nervous system: high (HF; 0.15-0.4 Hz), low (LF; 0.04-0.15 Hz), and very low frequency (VLF; 0.003-0.04 Hz). While the HF band represents parasympathetic activity, the LF band reflects both parasympathetic and sympathetic activity and is particularly related to blood pressure regulation. The VLF band is mainly related to the thermoregulation, vasomotor, and renin-angiotensin system. In the time domain, variation of IBIs can be quantified by different parameters such as the root mean square of successive differences between adjacent heartbeats (RMSSD), which is used to estimate variation mainly related to the HF band (pointing to parasympathetic activity; for other time-domain parameters, see [13]). By using non-linear methods (i.e., entropy domain), the complexity and irregularity of the IBI signal can be investigated. Approximate entropy (ApEn), detrended fluctuation analysis scaling exponent (DfaAlpha), Hurst exponent (Hurst), and sample entropy (SampEn) are amongst the most commonly used non-linear methods for HRV analysis [13,24,25]. Besides HRV, also heart rate (HR) provides information about autonomic nervous system regulation. Specifically, HR is a basic measure of overall cardiovascular arousal, which is influenced by both sympathetic activation and parasympathetic withdrawal.

## 2. Materials and Methods

### 2.1. Patients

From a total of 38 non-sedated and spontaneously breathing patients with DOC, nine patients had to be excluded from the analyses due to severe cardiac arrhythmias. Three further patients were excluded because they were exposed to constant 24-h light. Thus, 26 patients (8 women) between 16 and 80 years old (*mdn* = 51.61) with different etiologies (traumatic brain injury [TBI]: *n* = 11, non-traumatic brain injury [NTBI]: *n* = 15) from clinics in Austria (*n* = 12) and Belgium (*n* = 14) were included in the analyses. The patients’ behavioral state was assessed with the CRS-R [6]. While 16 patients were diagnosed with UWS, nine patients were in an MCS and one was in an EMCS. MCS and EMCS patients were combined for statistical analyses. Note that data of both patient samples (i.e., Austria and Belgium) have been used in two previous publications, where we studied diurnal variations in EEG parameters [26], as well as circadian variations in skin temperature [27] of patients with DOC. However, these studies did not focus on diurnal variations in electrocardiogram (ECG) data. The studies have been approved by the local ethics committees and informed consent was obtained from the patients’ legal representatives. For details on the study sample, see *Table 1*.

**Table 1.**
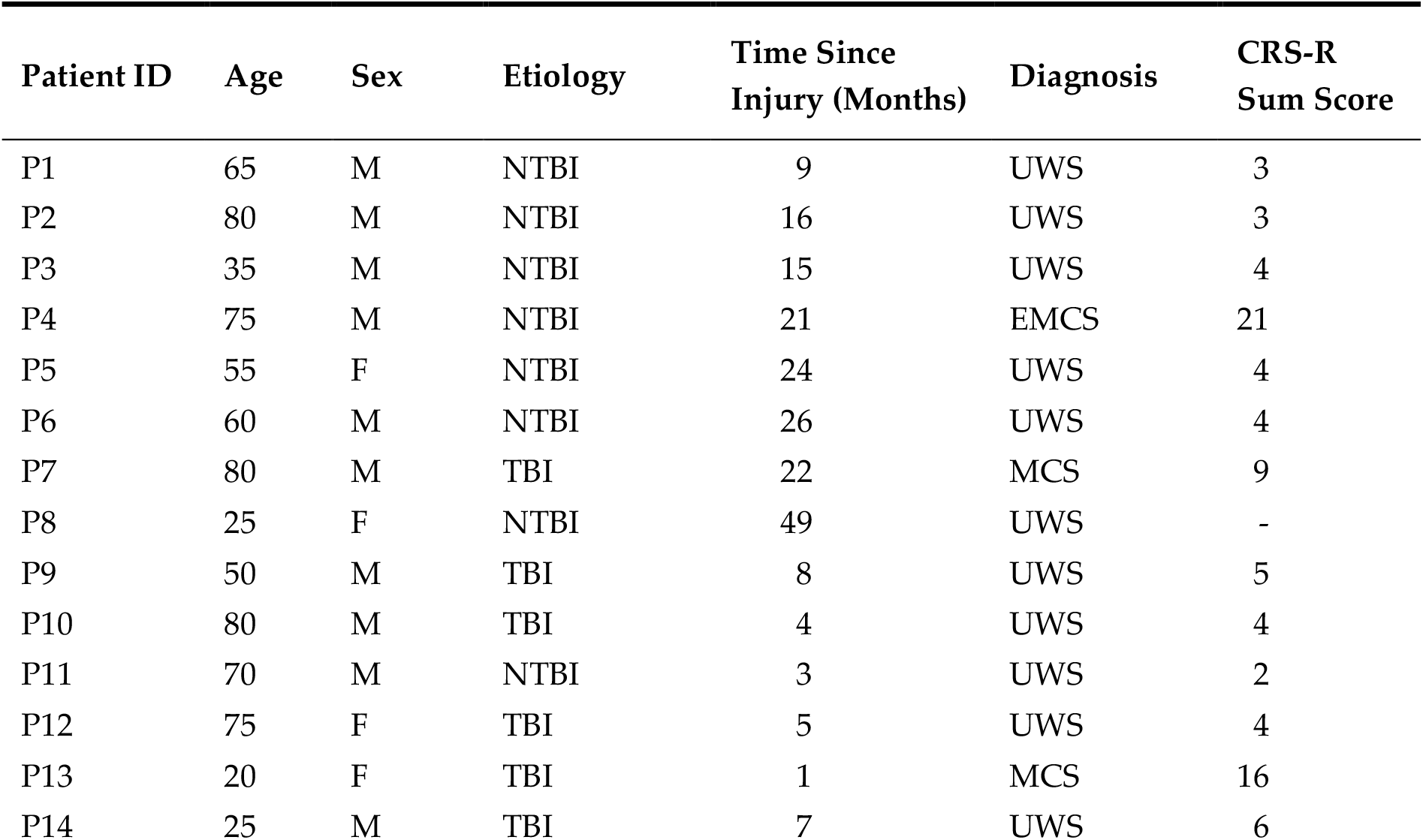
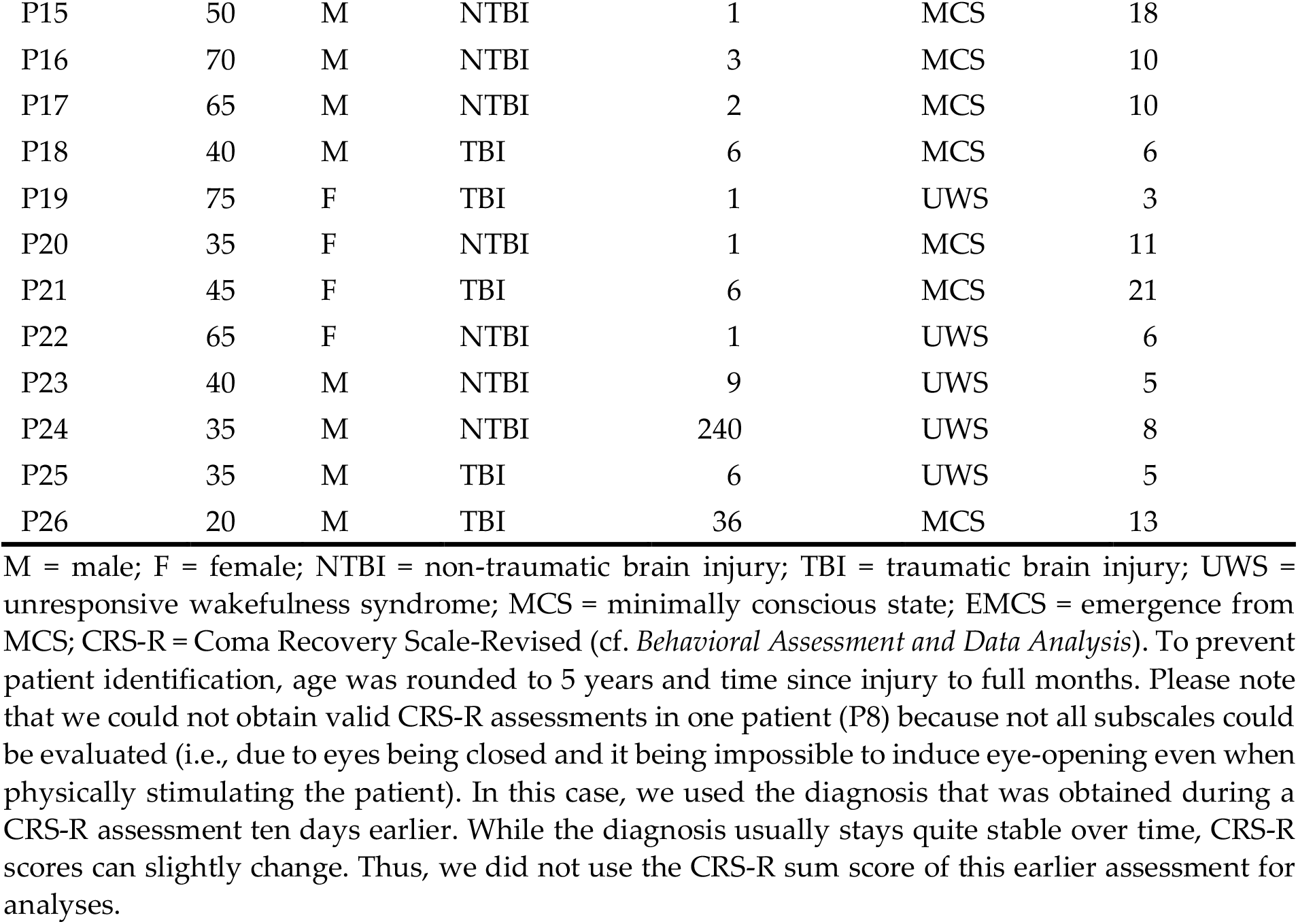
Demographic information and highest CRS-R sum score/diagnosis.

| Patient ID | Age | Sex | Etiology | Time Since Injury (Months) | Diagnosis | CRS-R Sum Score |
| --- | --- | --- | --- | --- | --- | --- |
| P1 | 65 | M | NTBI | 9 | UWS | 3 |
| P2 | 80 | M | NTBI | 16 | UWS | 3 |
| P3 | 35 | M | NTBI | 15 | UWS | 4 |
| P4 | 75 | M | NTBI | 21 | EMCS | 21 |
| P5 | 55 | F | NTBI | 24 | UWS | 4 |
| P6 | 60 | M | NTBI | 26 | UWS | 4 |
| P7 | 80 | M | TBI | 22 | MCS | 9 |
| P8 | 25 | F | NTBI | 49 | UWS | - |
| P9 | 50 | M | TBI | 8 | UWS | 5 |
| P10 | 80 | M | TBI | 4 | UWS | 4 |
| P11 | 70 | M | NTBI | 3 | UWS | 2 |
| P12 | 75 | F | TBI | 5 | UWS | 4 |
| P13 | 20 | F | TBI | 1 | MCS | 16 |
| P14 | 25 | M | TBI | 7 | UWS | 6 |
| P15 | 50 | M | NTBI | 1 | MCS | 18 |
| P16 | 70 | M | NTBI | 3 | MCS | 10 |
| P17 | 65 | M | NTBI | 2 | MCS | 10 |
| P18 | 40 | M | TBI | 6 | MCS | 6 |
| P19 | 75 | F | TBI | 1 | UWS | 3 |
| P20 | 35 | F | NTBI | 1 | MCS | 11 |
| P21 | 45 | F | TBI | 6 | MCS | 21 |
| P22 | 65 | F | NTBI | 1 | UWS | 6 |
| P23 | 40 | M | NTBI | 9 | UWS | 5 |
| P24 | 35 | M | NTBI | 240 | UWS | 8 |
| P25 | 35 | M | TBI | 6 | UWS | 5 |
| P26 | 20 | M | TBI | 36 | MCS | 13 |

M = male; F = female; NTBI = non-traumatic brain injury; TBI = traumatic brain injury; UWS = unresponsive wakefulness syndrome; MCS = minimally conscious state; EMCS = emergence from MCS; CRS-R = Coma Recovery Scale-Revised (cf. *Behavioral Assessment and Data Analysis*). To prevent patient identification, age was rounded to 5 years and time since injury to full months. Please note that we could not obtain valid CRS-R assessments in one patient (P8) because not all subscales could be evaluated (i.e., due to eyes being closed and it being impossible to induce eye-opening even when physically stimulating the patient). In this case, we used the diagnosis that was obtained during a CRS-R assessment ten days earlier. While the diagnosis usually stays quite stable over time, CRS-R scores can slightly change. Thus, we did not use the CRS-R sum score of this earlier assessment for analyses.

### 2.2. Study Protocol

#### 2.2.1. Austria

In Austria, the study protocol comprised two within-subject conditions, namely the (i) habitual light (HL) and (ii) dynamic daylight (DDL) condition. Each condition lasted one week and orders were randomized. While patients were in a room with standard clinic room lighting in the HL condition, patients were in a room with a ‘biodynamic’ room light in the DDL condition. During both weeks, ECG, skin temperature, and actimetry were assessed continuously. The patients’ behavioral repertoire was assessed twice with the CRS-R at the end of each week, once in the morning and once in the afternoon, in all patients (*n* = 12). The data from the DDL condition are beyond the scope of this study. For the current analyses, the CRS-R assessments from the HL condition as well as the 24-h ECG data recorded at the beginning of the HL condition are of interest and were analyzed (for more details on the study protocol, see [27]).

#### 2.2.1. Belgium

In Belgium, a 24-h polysomnography (PSG; including EEG, ECG, electromyogram [EMG], electrooculogram [EOG], and respiration) was performed in the patients’ usual clinical environment with standard clinic room lighting (i.e., HL). The CRS-R was performed twice (i.e., before and after the recording) in 9, and once in 5 of the 14 patients.

For the current analyses, the data of the Belgian patient sample was combined with the HL data of the Austrian patient sample, as lighting conditions were comparable in these two datasets. Thus, we are looking at HR/HRV under standard clinic room lighting conditions.

### 2.3. Behavioral Assessment and Data Analysis

#### 2.3.1. Coma Recovery Scale-Revised

The behavioral state of the patients was assessed with the CRS-R [6], which is composed of six subscales reflecting auditory, visual, motor, oromotor, communication and arousal functions with a total of 23 items. While the lowest threshold item on each subscale represents reflexive behavior, the highest threshold item indicates cognitively mediated behavior. The CRS-R is performed in a hierarchical manner, which means that the examiner starts with the highest item of each subscale and moves down the scale until the patient’s response meets the criteria for one item. When two CRS-R assessments were available, we used the CRS-R assessment where the patients showed the highest behavioral reactivity (i.e., the best diagnosis or highest sum score) as this is thought to best represent the ‘true’ state of the patient. The highest CRS-R score and diagnosis of each patient is shown in *Table 1*.

#### 2.3.2. Electrocardiography

For recording ECG in the patient sample from Austria, an ambulatory three-channel ECG device (eMotion Faros 180°, Mega Electronics Ltd, Kuopio, Finland) with self-adhesive Ambu^®^ BlueSensor SP electrodes (AMBU A/S, Ballerup, Denmark) was used. Two electrodes were placed in an infraclavicular position on the right and left body side, and one on a rib on the lower left thoracic wall. The sampling rate was 1,000 Hz.

In the patient sample from Belgium, the ECG was recorded as part of the PSG. For PSG recordings, BrainProducts amplifiers (BrainProducts, Gilching, Germany) were used. All PSG signals were recorded with a sampling rate of 500 Hz. For ECG recordings, two goldcup electrodes were used. One electrode was affixed in an infraclavicular position on the right body side and one on a rib on the lower left thoracic wall using EC2 electrode gel (Astro-Med^®^ Inc., West Warwick, USA).

#### 2.3.3. Heart Rate and Heart Rate Variability

To explore diurnal variations in HR/HRV, we used continuous 24-h ECG data from each patient, and divided the ECG recording into periods of clear daytime (i.e., forenoon: 8am-2pm, afternoon: 2pm-8pm) and nighttime (i.e., 11pm-5am) using BrainVision Analyzer 2.0 Software (Brain Products GmbH, Gilching, Germany). Importantly, all three periods were of equal length (i.e., 6 h). Periods with conditions of twilight (i.e., dawn: 5-8am, dusk: 8-11pm) were excluded from the analyses.

HRV analyses were conducted in ANSLAB 2.6 [28]. Accurate automatic R peak detections (obtained with the software’s algorithms) were carefully visually checked for the entire data of each patient and corrected whenever necessary. In a second and important preprocessing step, the IBI data was visually inspected for arrhythmias (e.g., ectopic beats) as single spikes in the IBI signal can seriously distort the spectral estimates of HRV [29]. While single spikes were interpolated with the software’s algorithm (i.e., mean of the adjacent IBIs), longer phases of arrhythmia were marked and excluded from the analyses (i.e., set to missing). In total, we removed per patient and daytime on average 0.3% (*min* = 0%, *max* = 7.2%) of the data points (for the amount of individual patients’ missing data, see *Supplementary material: Table S1*).

Patients’ HRV data were analyzed in the frequency, time, and entropy domains. Due to obvious non-stationarities in the IBI signals (i.e., sudden shifts in the mean and/or standard deviation) we used complex demodulation (CDM) for quantifying the HRV frequency parameters. CDM produces equivalent results to fast Fourier transform but is less affected by non-stationarity (a prerequisite for accurate fast Fourier transform) [30,31].

We computed the mean IBI oscillation amplitude (in ms) for the very low (VLF; 0.003-0.04 Hz), low (LF; 0.04-0.15 Hz), and high (HF; 0.15-0.4 Hz) frequency bands for each minute. During the export of the mean IBI oscillation amplitude, ANSLAB automatically interpolates segments that were set to missing. Thus, we post-hoc excluded all segments that were automatically interpolated using R version 3.6.1. [32]. Mean IBI (in ms; i.e., the time interval between two successive R-peaks), mean HR (equivalent to 60,000/IBI; in beats/min), and mean RMSSD (in ms) were also computed for each minute. For statistical analyses of HR/HRV parameters, all one-minute segments within each 6-h period (i.e., 8am-2pm, 2pm-8pm, and 11pm-5am) were averaged using R. Thus, we arrived at one value per patient and HR/HRV parameter for forenoon, afternoon, and night. For quantifying the complexity of the patients’ HR variations, approximate entropy (ApEn), detrended fluctuation analysis scaling exponent (DfaAlpha), sample entropy (SampEn), and the Hurst exponent (Hurst) were computed for the whole 6-h segments (i.e., forenoon, afternoon, and night). ApEn describes the likelihood that patterns remain similar for subsequent comparisons, with higher ApEn values indicating higher irregularity and complexity in time-series data. DfaAlpha quantifies the presence or absence of fractal correlation properties in non-stationary time-series data, with higher DfaAlpha values suggesting fractal-like HR dynamics. SampEn describes the probability that two sequences of 1-5 consecutive data points (i.e., SampEn1-5) that are similar to each other will remain similar when one consecutive point is included. Higher SampEn indicates frequent incidence of dissimilarities in the time-series data. The Hurst exponent is a quantification of long-range dependence, with values of 0.5-1 indicating a long-term positive correlation meaning that a high value in the series will probably be followed by another high value, values of 0-0.5 indicating time series with long-term negative correlation or switching between high and low values, and a value of 0.5 suggesting that the series are uncorrelated. For more details on non-linear HRV analyses, see [24,25].

#### 2.3.4. Respiration

Since DOC may be related to respiratory alterations [33] and as low respiration rates (i.e., <9 cycles per minute [cpm] = <0.15 Hz) can affect the HF estimates of HRV via shifts in the respiratory sinus arrhythmia to the LF band [34], we analyzed the respiratory signal in a subsample of patients (*n* = 12) for whom respiration was simultaneously recorded via a respiration belt around the thorax. Mean respiratory rate for each minute was computed in ANSLAB and averaged for 5-minute segments using R. We found that only one 5-min segment (i.e., 1.39% of 6-h recordings) of two patients (i.e., P17: forenoon; P26: night) was <9 cpm. Thus, we decided that there was no need to control the HF parameter for respiration in our patient sample. Although respiration data was only available from a subset of patients, it is unlikely that breathing patterns in the other patients were appreciably different.

#### 2.3.4. EEG Permutation entropy

For analyzing heart-brain interaction, we correlated HRV with EEG entropy in the Belgian patient sample (*n* = 14) where PSG was recorded. As the EEG data of the Belgian sample was already used in a previous study, where EEG permutation entropy (PE) had been computed, we used the EEG entropy values of the respective patients from that publication [26]. More specifically, PE of the entire EEG signal was computed for day (i.e., 8am-8pm) and night (i.e., 11pm-5am). PE quantifies the level of irregularity or unpredictability of an EEG signal. Higher PE values indicate more complex and/or random signals. For further information on the preprocessing of the EEG signal and the entropy analyses, see [26].

### 2.4. Statistical Analyses

Statistical analyses were done in R. As not every variable was normally distributed (i.e., Shapiro-Wilk test for normality: *p* < .001; cf. *Supplementary material: Table S2*), we used an advanced semi-parametrical statistical approach. The significance level was set to α = .05 (two-sided) for all analyses. As suggested by Wasserstein et al. [35], we interpreted the overall pattern rather than focusing on individual *p*-values. Therefore, we also interpreted *p*-values .05 < *p* ≤ .10 if they were in line with the overall pattern of results, and also ‘credible’ from a Bayesian point of view. More specifically, we additionally tested all trends of our post-hoc comparisons and correlations analyses with Bayesian multilevel regression models via the Stan-based ‘brms’ package available for R [36,37]. We report regression coefficients and the 95% credible intervals (CIs; i.e., Bayesian confidence intervals). The CI describes the interval in which a parameter value falls with a 95% probability given the data observed, prior and model assumptions. Thus, an effect is considered to significantly differ from zero if zero is not included in the CI. We used weak- or non-informative default priors whose influence on results is negligible. For all computed regression models no divergent transitions occur, Rhat (i.e., potential scale reduction factor on split chains) was < 1.01, and effective sample size (ESS) was > 400.

For the analyses of differences in HR/HRV parameters (i.e., IBI, HR, RMSSD, VLF, LF, HF, ApEn, DfaAlpha, Hurst, and SampEn) and EEG entropy (i.e., PE) between different times of the day (i.e., within-subjects factor; forenoon, afternoon, night), diagnoses [i.e., between-subjects factor; (E)MCS vs. UWS], etiology [i.e., between-subjects factor; TBI vs. NTBI], or sex [i.e., between-subjects factor; female vs. male], we used advanced non-parametric analyses for repeated measures designs as implemented in the ‘MANOVA.RM’ package available for R [38]. We report the resampled Wald-type statistic (WTS). As a resampling method, the function ‘perm’ was used, which randomly permutes all observations. The number of iterations used to calculate the resampled statistic was 10,000. To correct for multiple tests, *p*-values of post hoc comparisons were adjusted using the method of Benjamini and Hochberg (BH) [39] as implemented in the ‘p.adjust’ function in R. For correlation analyses of HR/HRV parameters, CRS-R sum score, and EEG entropy we report Kendall’s Tau.

## 3. Results

### 3.1. Interbeat Interval and Heart Rate

Analyses of the IBIs of 26 patients revealed a trend towards a main effect for *time* (*F*_*WTS*_(2)=6.52, *p*=.068) and a significant effect for *diagnosis* (*F*_*WTS*_(1)=5.8, *p*=.028), but no significant *time* × *diagnosis* interaction (*F*_*WTS*_(2)=1.36, *p*=.523). Specifically, patients showed larger IBIs during the night as compared to forenoon (*F*_*WTS*_(1)=7.76, *p*=.027) and afternoon (*F*_*WTS*_(1)=4.74, *p*=.059; *b*=-37.44, 95%CI=[-67.46, -7.42]). No differences could be observed in the patients’ IBIs between fore- and afternoon (*F*_*WTS*_(1)=0.21, *p*=.648; cf. *Figure 1a*). Further, patients in UWS showed larger IBIs as compared to patients in (E)MCS (*F*_*WTS*_(1)=5.8, *p*=.028; cf. *Figure 1b*).

**Figure 1.**
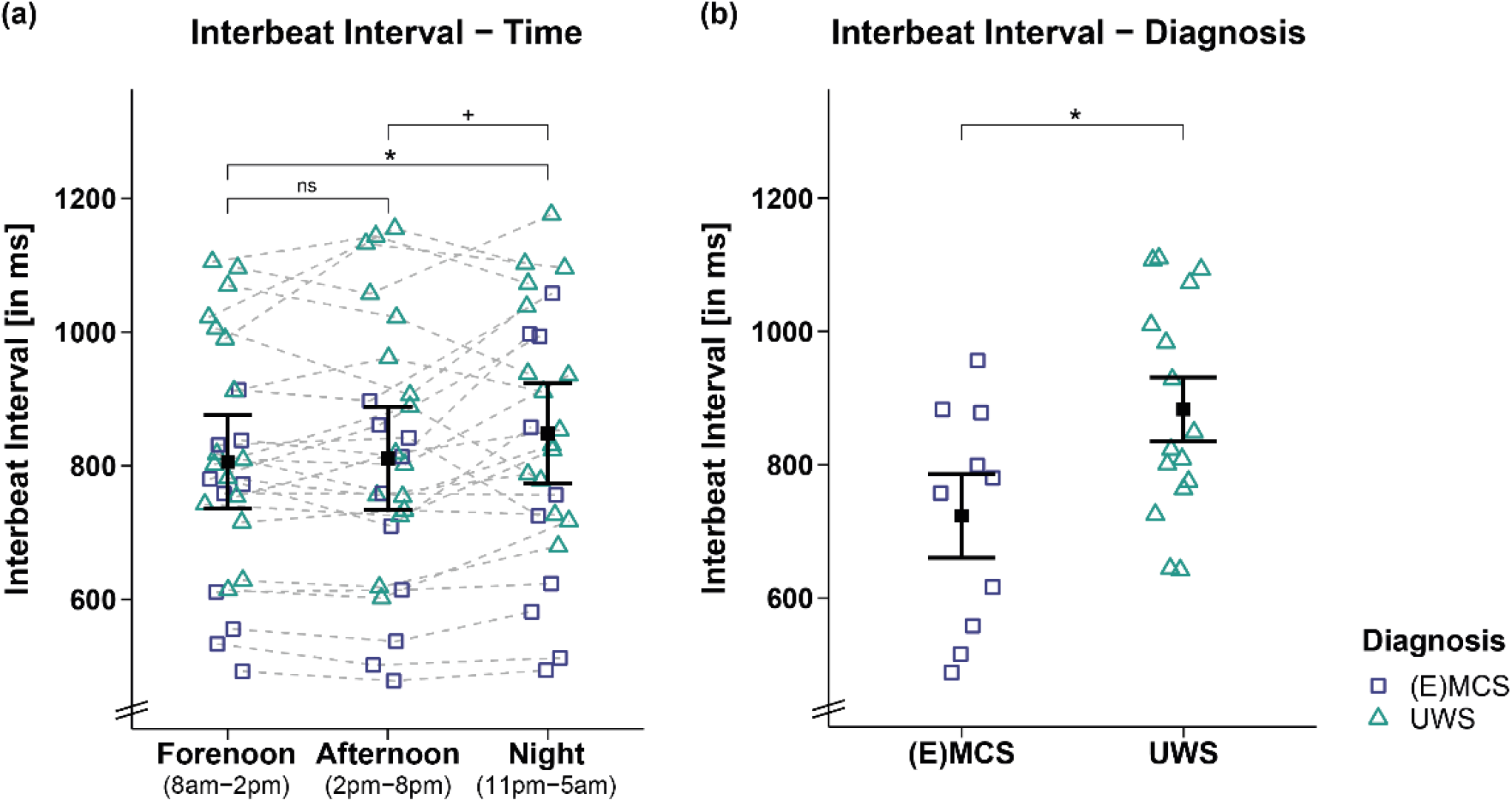
Interbeat interval (IBI) separately for time and diagnosis contrasts. **(a)** While patients’ IBIs were larger during the night as compared to the day (i.e., forenoon, afternoon), they did not differ between fore- and afternoon.**(b)** Patients in UWS showed larger IBIs than patients in (E)MCS. Error bars represent the mean and 95% confidence interval. \**p* < .05, ^+^*p* ≤ .1, ns = not significant. Abbreviations: (E)MCS = (emergence from) minimally conscious state, UWS = unresponsive wakefulness syndrome, ms = milliseconds.

This effect was supported by correlation analyses of IBIs and CRS-R sum scores of 25 patients, showing that lower CRS-R sum scores were associated with larger IBIs during fore- (*r*τ(23)=-0.34, *p*=.02; cf. *Figure 2a*) and afternoon (*r*τ(23)=-0.38, *p*=.009; cf. *Figure 2b*) and by trend during the night (*r*τ(23)=-0.27, *p*=.07; *cf. Figure 2c*). However, we refrain from interpreting the effect during the night as it is does not appear robust from a Bayesian point of view (*b*=-12.34, 95%CI=[-25.79, 1.16]).

**Figure 2.**
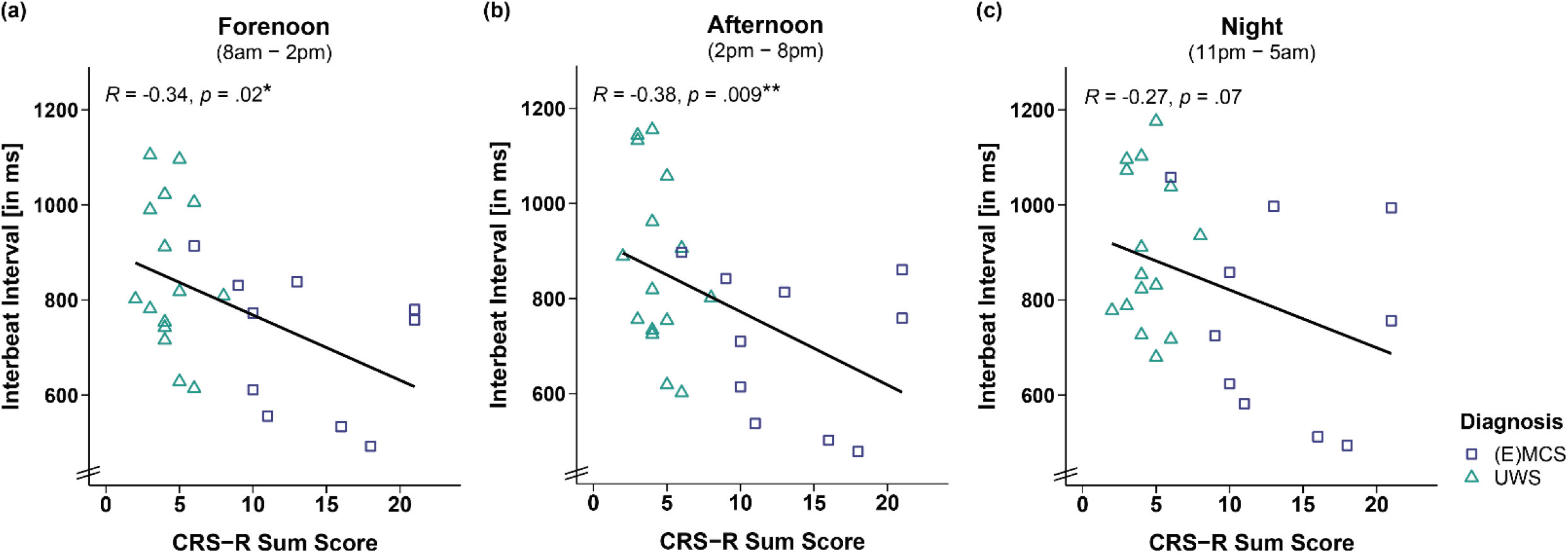
Correlation between interbeat interval (IBI) and CRS-R sum score separately for time. Larger IBIs were associated with lower CRS-R sum scores throughout the **(a, b)** day (i.e., forenoon, afternoon) and **(c)** night. Please note that the effect during the night is no longer ‘credible’ from a Bayesian point of view, and will not be interpreted.\*\**p* < .01, \**p* < .05. Abbreviations: (E)MCS = (emergence from) minimally conscious state, UWS = unresponsive wakefulness syndrome, ms = milliseconds.

Analyses of HR yielded similar results (cf. *Supplementary material: Figures S1 and S2*), which is expected as HR is inverse proportional to the IBI signal. In both, IBI and HR, no differences for etiology (i.e., TBI vs. NTBI) or sex (i.e., male vs. female) were observed (cf. *Supplementary material: Tables S3 and S4*).

### 3.2. HRV Time Domain

Analyses of the RMSSD of 26 patients did not reveal significant main effects for *time* (*F*_*WTS*_(2)=0.4, *p*=.828), *diagnosis* (*F*_*WTS*_(1)=0.37, *p*=.554) and *time* × *diagnosis* interaction (*F*_*WTS*_(2)=3.91, *p*=.174; cf. *Figure 3*). No differences for etiology (i.e., TBI vs. NTBI) or sex (i.e., male vs. female) were observed (cf. *Supplementary material: Tables S3 and S4*).

**Figure 3.**
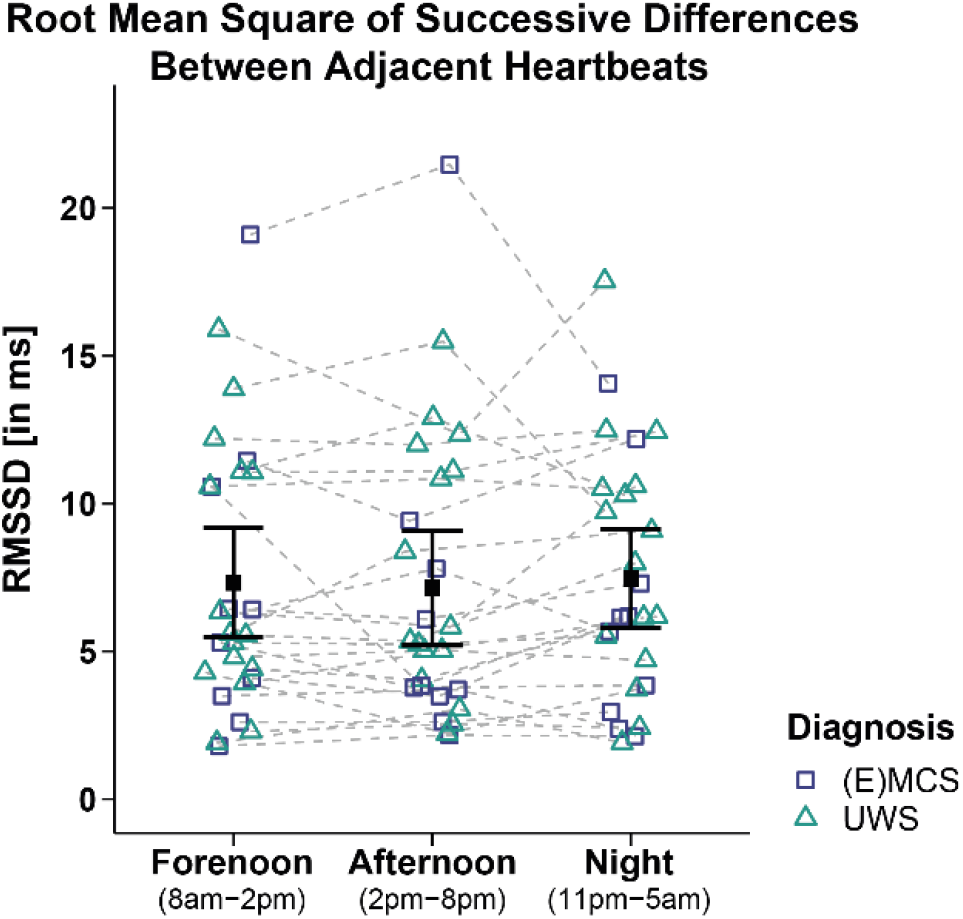
Root mean square of successive differences between adjacent heartbeats (RMSSD). Patients’ RMSSD did not differ between times and diagnoses. Error bars represent the mean and 95% confidence interval. Abbreviations: (E)MCS = (emergence from) minimally conscious state, UWS = unresponsive wakefulness syndrome, ms = milliseconds.

### 3.3. HRV Frequency Domain

Analyses of VLF of 26 patients yielded a trend towards a main effect for *time* (*F*_*WTS*_(2)=6.63, *p*=.064), but no significant effect for *diagnosis* (*F*_*WTS*_(1)=1.74, *p*=.207) and the *time* × *diagnosis* interaction (*F*_*WTS*_(2)=1.08, *p*=.603). Post hoc comparisons of VLF between times of day did not yield significance anymore after correcting for multiple comparisons (i.e., fore- vs. afternoon: *F*_*WTS*_(1)=1.15, *p*=.295; forenoon vs. night: *F*_*WTS*_(1)=1.26, *p*=.295; afternoon vs. night: *F*_*WTS*_(1)=4.73, *p*=.111 [before BH-correction: *p*=.03]; cf. *Figure 4a*).

**Figure 4.**
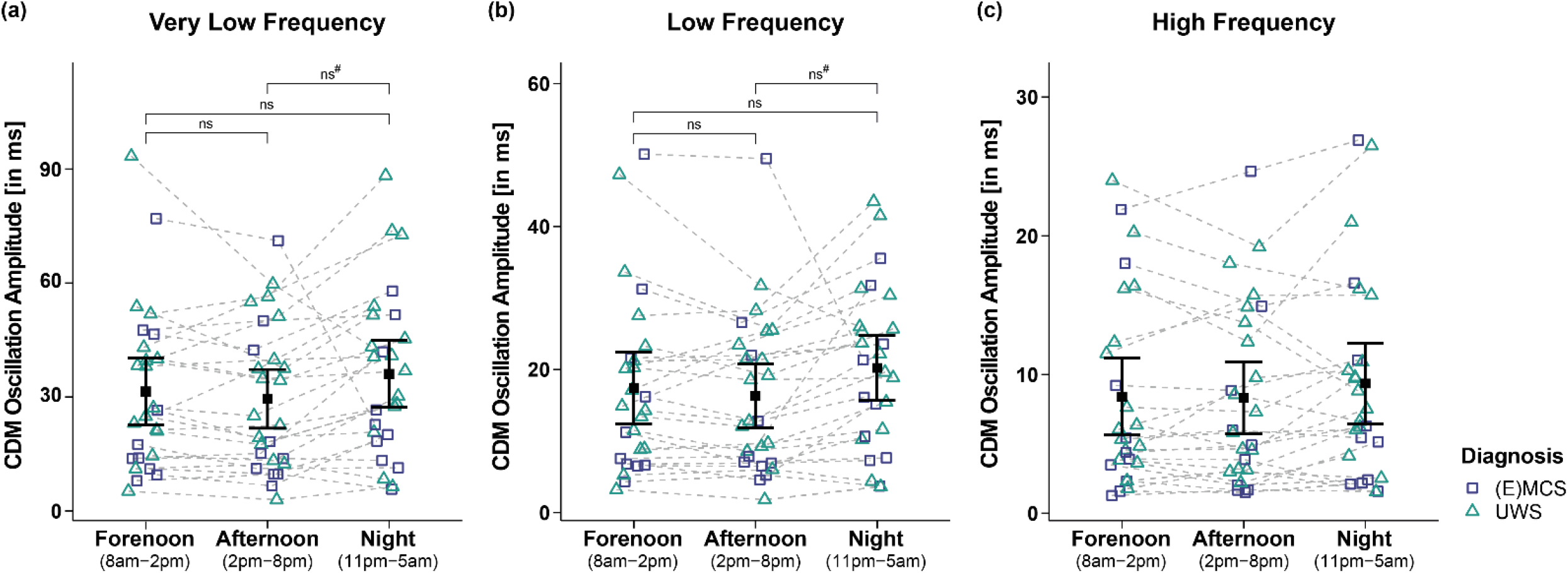
Very low (VLF), low (LF), and high frequency (HF) oscillation amplitude of the IBI signal computed by complex demodulation (CDM, see *Heart Rate and Heart Rate Variability* for details). Analyses of the **(a)** VLF and **(b)** LF band revealed significant main effects for time (i.e., forenoon, afternoon, night). However, post-hoc comparisons between times of day did not yield significance anymore after correction for multiple comparisons. **(c)** Patients’ HF did also not differ between times and diagnoses. Error bars represent the mean and 95% confidence interval. # = significant before correction for multiple comparisons, ns = not significant. Abbreviations: (E)MCS = (emergence from) minimally conscious state, UWS = unresponsive wakefulness syndrome, CDM = complex demodulation, ms= milliseconds.

Analyses of LF of 26 patients revealed a trend towards a main effect for *time* (*F*_*WTS*_(2)=5.69, *p*=.088), but no significant effect for *diagnosis* (*F*_*WTS*_(1)=0.46, *p*=.51) and *time* × *diagnosis* interaction (*F*_*WTS*_(2)=0.36, *p*=.843). Post hoc comparisons of LF between times of day did not yield significance anymore after correcting for multiple comparisons (i.e., forenoon vs. afternoon: *F*_*WTS*_(1)=1.65, *p*=.213; forenoon vs. night: *F*_*WTS*_(1)=1.75, *p*=.213; afternoon vs. night: *F*_*WTS*_(1)=4.86, *p*=.108 [before BH-correction: *p*=.028]; cf. *Figure 4b*).

Analyses of HF of 26 patients did not reveal significant main effects for *time* (*F*_*WTS*_(2)=3.8, *p*=.191), *diagnosis* (*F*_*WTS*_(1)=0.57, *p*=.460), and *time* × *diagnosis* interaction (*F*_*WTS*_(2)=0.04, *p*=.980; cf. *Figure 4c*).

No etiology or sex differences were observed for any frequency band (cf. *Supplementary material: Tables S3 and S4*).

### 3.4. HRV Entropy Domain

Analyses of ApEn of 26 patients revealed a significant main effect for *time* (*F*_*WTS*_(2)=21.35, *p*=.001), but no significant effect for *diagnosis* (*F*_*WTS*_(1)=1.79, *p*=.197) and the *time* × *diagnosis* interaction (*F*_*WTS*_(2)=1.99, *p*=.395). Specifically, patients showed a higher ApEn during forenoon as compared to afternoon (*F*_*WTS*_(1)=15.79, *p*<.001). No differences could be observed in the patients’ ApEn during night as compared to forenoon (*F*_*WTS*_(1)=2.19, *p*=.154) and afternoon (*F*_*WTS*_(1)=2.77, *p*=.154; cf. *Figure 5*). Analyses of other entropy parameters did not yield significant main effects for *time* and *diagnoses*, and the *time* × *diagnosis* interaction (cf. *Supplementary material: Table S5*).

**Figure 5.**
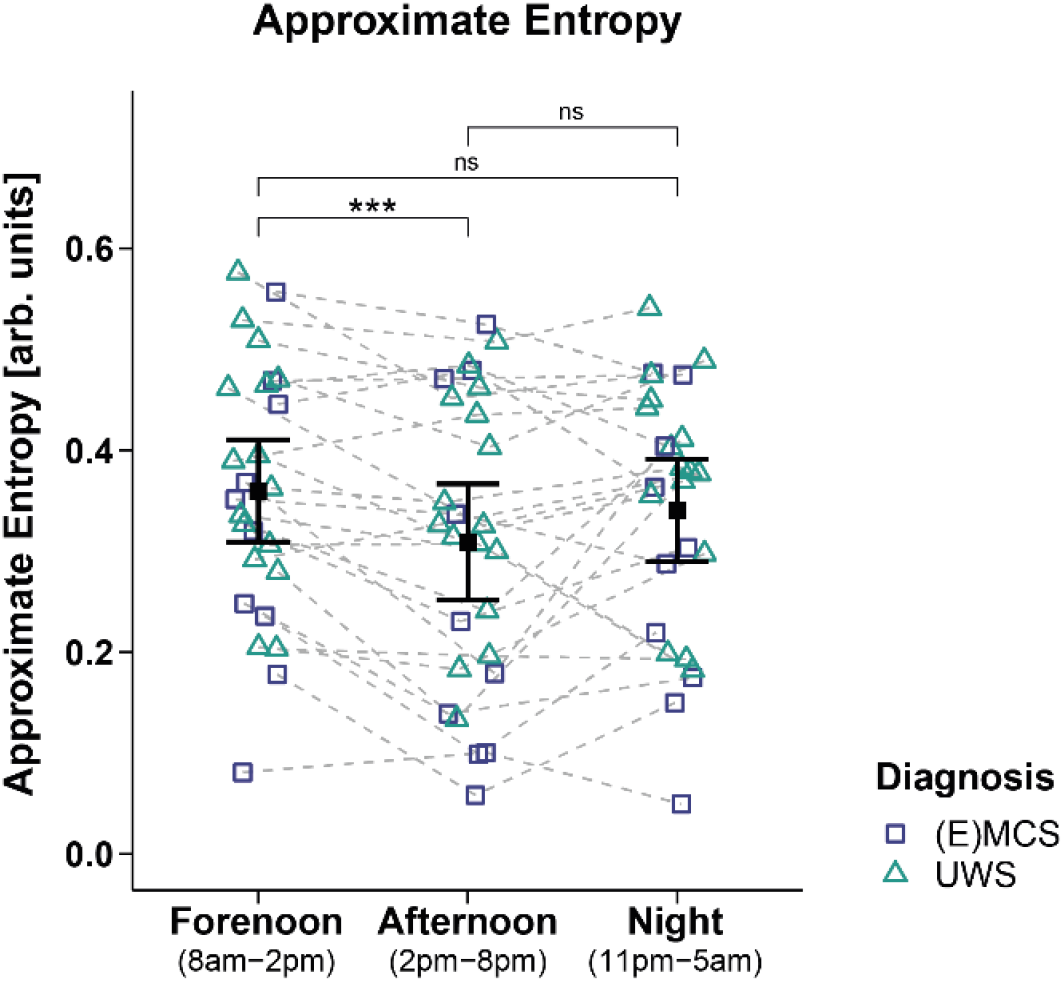
Approximate entropy (ApEn). Patients showed a higher ApEn during forenoon as compared to afternoon. No significant differences were evident between night and day (i.e., night vs. forenoon or afternoon). Error bars represent the mean and 95% confidence interval. \*\*\**p* < .001, ns = not significant. Abbreviations: (E)MCS = (emergence from) minimally conscious state, UWS = unresponsive wakefulness syndrome, arb. units = arbitrary units.

However, analyses of the DfaAlpha and SampEn1 yielded a significant main effect for etiology, with higher DfaAlpha (*F*_*WTS*_(1)=4.59, *p*=.043; cf. *Figure 6a*) and SampEn1 (*F*_*WTS*_(1)=4.36, *p*=.049; cf. *Figure 6b*) being observed in patients with TBI as compared to patients with NTBI. No etiology and sex differences were observed for the other entropy parameters (cf. *Supplementary material: Tables S3 and S4*).

**Figure 6.**
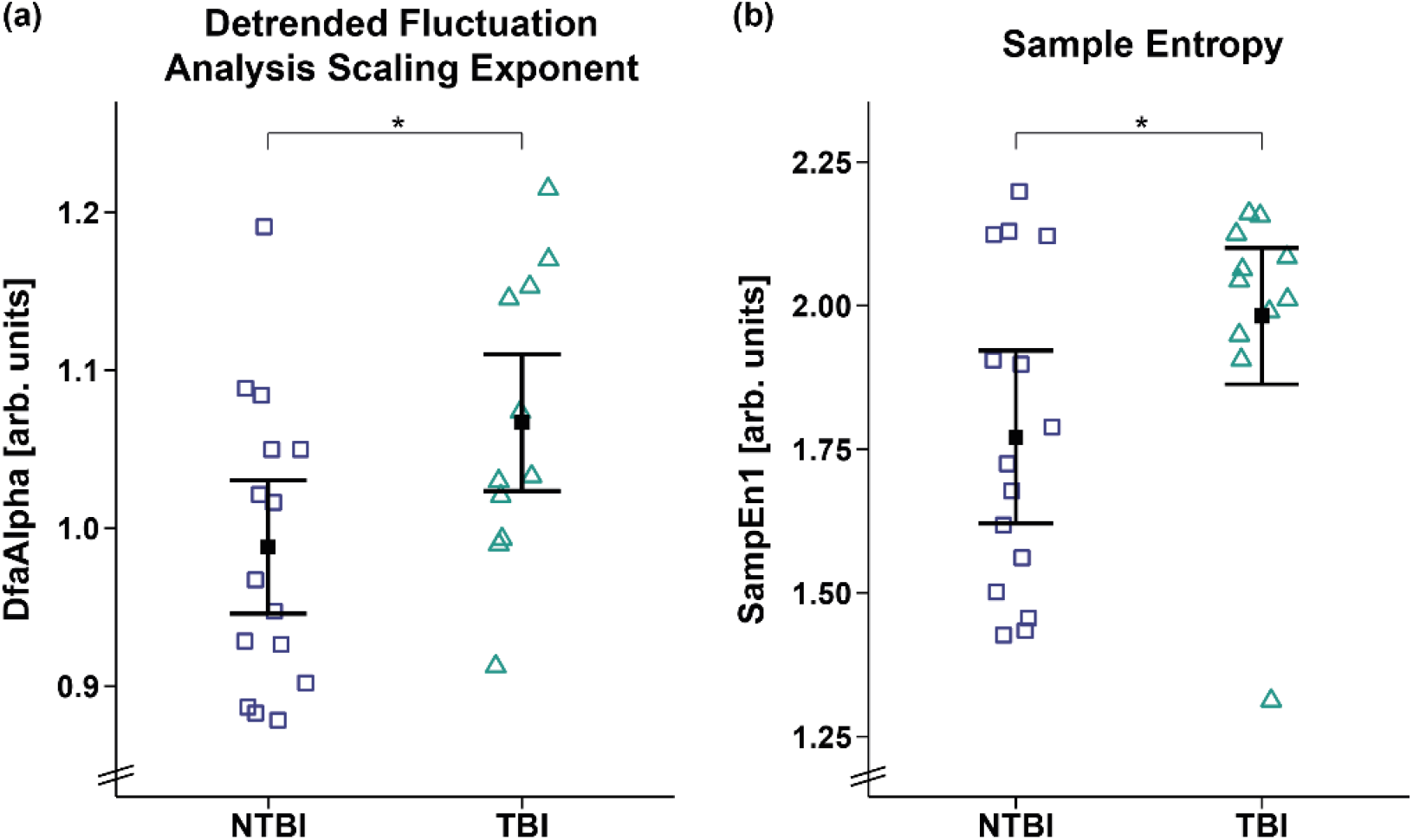
Detrended fluctuation analysis scaling exponent (DfaAlpha) and sample entropy 1 (SampEn1). Patients with TBI showed a higher **(a)** DfaAlpha and **(b)** SampEn1 as compared to patients with NTBI. Error bars represent the mean and 95% confidence interval. \**p* < .05. Abbreviations: NTBI = non-traumatic brain injury, TBI = traumatic brain injury, arb. units = arbitrary units.

### 3.5. Correlation of EEG and HRV Entropy

Correlation analyses in 14 patients showed that a higher ApEn was associated with a higher PE during the night (i.e., 11pm-5am; *F*_*WTS*_(1)=0.47, *p*=.019; cf. *Figure 7b*). No significant correlation was observed during the day (i.e., 8am-8pm, *r*τ(12)=0.3, *p*=.157, cf. *Figure 7a*).

**Figure 7.**
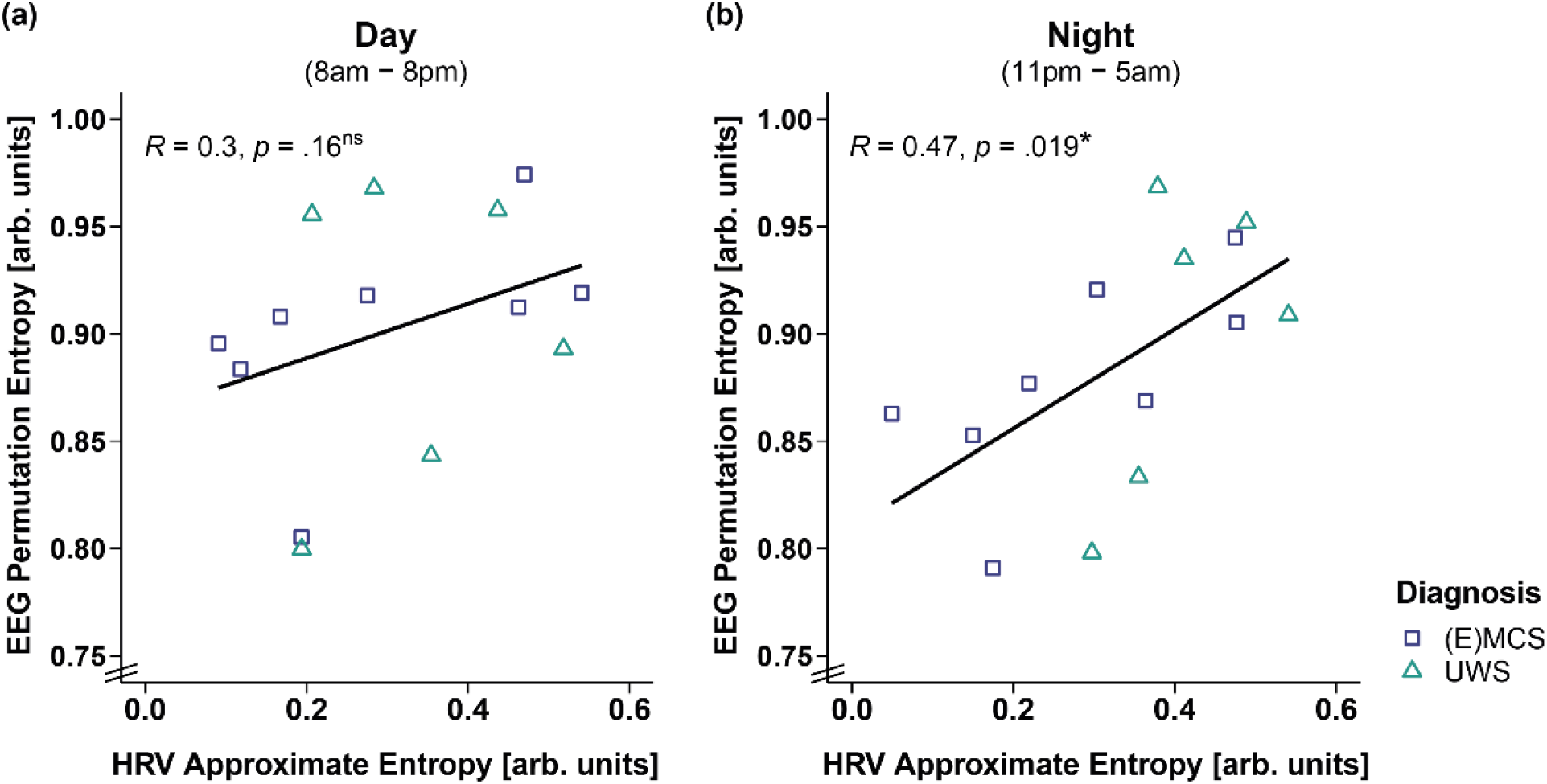
Correlation between EEG permutation entropy (PE) and HRV approximate entropy (ApEn) separately for day and night. **(a)** While EEG PE and HRV ApEn did not correlate during the day (i.e., 8am-8pm), **(b)** a higher EEG PE was associated with a higher HRV ApEn during the night (i.e., 11pm-5am). \**p* < .05, ns = not significant. Abbreviations: (E)MCS = (emergence from) minimally conscious state, UWS = unresponsive wakefulness syndrome, arb. units = arbitrary units.

## 4. Discussion

In patients with DOC, variations in cardiac activity show a diurnal pattern. Specifically, we find preserved diurnal variations in the length of the patients’ IBIs. IBIs were larger during the night than during the day indicating that, as in healthy individuals [20], the heart slows down during the night due to parasympathetic dominance reflecting relaxation and sleep. Further, the complexity of patients’ HRV signal varies across wakefulness, with the signal being more irregular and complex (i.e., higher ApEn) during forenoon as compared to afternoon. This has also been found in healthy individuals [40], and might be due to an increased cardiac sympathovagal response in the morning after awakening. More specifically, it has been shown in healthy individuals that sleep-to-wake transitions in the morning are associated with higher sympathetic activation compared to those occurring during the rest of the day [20]. Thus, although patients with DOC often fluctuate between sleep and wake-like phases – hence experiencing several ‘awakenings’ throughout the day – results indicate that the awakening in the morning is probably the one associated with the most prominent change in arousal. Further, it might also be the case that patients show less frequent or temporally more regular (i.e., systematic) alterations of arousal and/or awareness in the afternoon, which may be associated with lower entropy values. This would go in line with our findings from a previous study, where patients tended to exhibit higher CRS-R sum scores at a later daytime (i.e., afternoon) [7], which also requires more stable arousal and awareness levels. Another explanation for the entropy drop from fore- to afternoon might be the recovery from stressful events that possibly took place more frequently during forenoon (e.g., therapies, medical rounds, nursing activities). It has been shown in healthy controls that a stressful task leads to a significant reduction in entropy in the succeeding relaxation period [41]. Thus, the variation of cardiac activity over the day might be necessary for an optimal interaction and adaptation to changing demands in the environment. Interestingly, while we found that patients in UWS generally had larger IBIs (i.e., lower HRs) than patients in (E)MCS, lower IBIs were associated with higher behavioral reactivity (i.e., higher CRS-R sum scores) in patients with DOC only during the day, but not during the night. Looking at the data, one can see that this effect is mainly driven by the nocturnal slowing of the heart in patients in an (E)MCS, underlining that cardiac activity shows a diurnal pattern, particularly when patients have (partially) regained consciousness. Thus, our findings complement earlier research suggesting that better circadian rhythm integrity is associated with higher consciousness levels [7,21,22]. More specifically, we found in earlier studies that variations of peripheral biosignals, such as skin temperature, melatonin(-sulfate) and wrist actimetry, are better aligned to a healthy 24-h rhythm (i.e., circadian rhythm) and more pronounced in patients with a higher behavioral repertoire [7,21,22].

Concerning central biosignals, such as brain activity derived from EEG recordings, earlier studies have shown that more severely affected patients with UWS do not only show a stronger general slowing of the EEG, but also no clear diurnal pattern [26,42]. This is in line with findings we present here (i.e., lower HR/larger IBIs in patients with UWS) and possibly reflects the interaction between peripheral and central biosignals. Thus, we additionally investigated whether there is an association between the heart and central measures of brain activity, that is, EEG entropy (i.e., a measure describing the level of irregularity or unpredictability of the brain signal). We found that heart and brain activity are coupled during the night, but not during the day. Specifically, while during night, a higher EEG entropy (i.e., PE) was associated with a higher ECG entropy (i.e., ApEn), no such association was evident during the day. It might be the case that during the night (i.e., habitual sleep), and due to less disturbance by external cues from the environment and a stronger focus on internal processes, brain and body rhythms are better connected. Conversely, during the day, the brain focuses more on the processing of sensory signals in the environment, leading to a decrease in the synchrony between brain and body rhythms. Interestingly, the effect in our data seems to be mainly driven by EEG entropy. Specifically, patients showed lower EEG entropy values during the night as compared to the day (see *Supplementary material: EEG Entropy*), which might be an effect of the dominance of slow oscillations during the night (i.e., more synchronized brain activity, and thus less signal complexity) and probably indicates the existence of a ‘sleep-like state’ during the night. ECG entropy, however, does not differ between day and night. A reason for this could be that the patients spend most of the time in a lying position or a position where the upper body is raised in a 45° angle, which reduces events that usually influence HRV such as changes of posture or physical movements. The brain signal however, can still change independently – based on the patients’ clinical state. Specifically, while patients in an (E)MCS showed higher EEG complexity during the day than during the night, no diurnal variation was evident in patients with UWS (see *Supplementary material: EEG Entropy*).

Additionally, cardiac activity does not only inform about the integrity of diurnal variations and the degree of behavioral reactivity in patients with DOC, but also differentiates between patients’ etiologies. Specifically, patients with TBI showed a higher DfaAlpha and SampEn1 than patients with NTBI, suggesting fractal-like (i.e., aperiodic) HR dynamics and higher dissimilarities in the HRV signal of patients with TBI. In other words, cardiac activity is less complex and variable in patients with NTBI. Reduced HRV complexity has been shown to be a predictor for mortality [43,44]. This is in line with previous findings showing that patients with NTBI often have a less favorable prognosis than patients with TBI [45-47].

When looking at the time (i.e., RMSSD) and frequency (i.e., VLF, LF, HF) domains of HRV, no differences between time of day, diagnoses and etiologies were evident. One reason might be of methodological nature. More specifically, the activity of the heart is not regular/periodic, but rather fluctuates in complex/aperiodic patterns. Thus, it has frequently been argued that non-linear measures (i.e., measures of mathematical chaos/entropy) might be more appropriate for the analysis of HRV data [48,49]. This is in line with findings from a study that showed a difference in HRV entropy between patients with UWS and healthy individuals (i.e., lower ApEn in patients with UWS), but no such differences in any of the linear parameters (i.e., IBI, SDRR, RMSSD, LF/HF ratio) [17].

## 5. Conclusions

To summarize, patients with severe brain injuries – particularly those who (partially) regained consciousness – still had preserved diurnal variations as characterized by a heart rate slowing during the night. This suggests preserved integrity of circadian rhythms in the autonomic nervous system activity. Further, cardiac activity differentiated between patients’ etiologies and diagnoses. Patients with UWS had larger IBIs (i.e., lower heart rate) than patients in an (E)MCS, and patients with NTBI had a less complex HRV signal than patients with TBI. Thus, cardiac activity and its variations might represent a peripheral window to central (brain) functioning. Indeed, we found an interaction of heart and brain signal complexity, which also followed a diurnal pattern. Specifically, while a more complex brain signal was associated with a more complex heart signal during the night, no such association was found during the day. In conclusion, HR and HRV seem to mirror the integrity of brain functioning and consequently might serve as supplementary measures that aid the differentiation between clinical states. Ultimately, this has the potential to improve the validity of assessments in patients with DOC.

## Data Availability

The data that support the findings of this study are available from the corresponding authors upon reasonable request.

## Author Contributions

Conceptualization, Ma.S., C.B. and M.A.; methodology, Ma.S., M.A., C.B., M.L. and F.W.; software, M.A., M.L. and F.W.; formal analysis, M.A.; investigation, Ma.S., G.P., B.A. and Mo.S.; resources, Ma.S., F.W. and M.L.; data curation, M.A. and Ma.S..; writing—original draft preparation, M.A. and C.B.; writing—review and editing, M.A., C.B., F.W., Ma.S., M.L.; visualization, M.A.; supervision, Ma.S.; project administration, Ma.S. and M.A.; funding acquisition, Ma.S. All authors have read and agreed to the published version of the manuscript.

## Institutional Review Board Statement

The study was conducted according to the guidelines of the Declaration of Helsinki, and approved by the Ethics Committees of the Medical University of Liège and Graz.

## Informed Consent Statement

Informed consent was obtained from the legal representatives of all patients involved in the study.

## Acknowledgments

We thank Sarah Haberl, Kerstin Dokter, Marlen Dido Grand and Julius Köppen as well as the staff at the Hospital in Graz for their constant support and help with the data collection. Further, we thank Marina Meier and Jasmin Preiß for their support with the data preparation. Additionally, we thank Wislowska Malgorzata for sharing patients’ EEG entropy data with us. Finally, we also thank the Coma Science Group (Liège) for contributing data and the patients’ families for their trust and cooperation in this study.

## Trial Registration Information

German Clinical Trials Register (DRKS00016041); registration: 18.01.2019 https://www.drks.de/drks_web/navigate.do?navigationId=trial.HTML&TRIAL_ID=DRKS00016041

## Abbreviations

ApEn: Approximate entropy
BH: Benjamini-Hochberg correction for multiple comparisons
CDM: Complex demodulation
CI: Credibility interval
Cpm: Cycles per minute
CRS-R: Coma Recovery Scale – Revised
DDL: Dynamic daylight
DfaAlpha: Detrended fluctuation analysis scaling exponent
DOC: Disorders of consciousness
ECG: Electrocardiography
EEG: Electroencephalography
EMCS: Emergence from minimally conscious state
EMG: Electromyography
EOG: Electrooculography
fMRI: Functional magnetic resonance imaging
HF: High frequencies (0.15-0.4 Hz)
HL: Habitual light HR Heart rate
HRV: Heart rate variability Hurst Hurst exponent
IBI: Interbeat interval
LF: Low frequencies (0.04-0.15 Hz)
LF/HF ratio: Ratio between low and high frequencies
MCS: Minimally conscious state
NTBI: Non-traumatic brain injury
PE: Permutation entropy
PSG: Polysomnography
RMSSD: Root mean square of successive differences between adjacent heartbeats SampEn Sample entropy
SDRR: Standard deviation of interbeat intervals
TBI: Traumatic brain injury
UWS: Unresponsive wakefulness syndrome
VLF: Very low frequencies (0.003-0.04 Hz)
WTS: Wald-type statistic

## References

1. Laureys, S. The neural correlate of (un)awareness: lessons from the vegetative state. Trends Cogn Sci 2005, 9, 556–559, doi:10.1016/j.tics.2005.10.010.

2. Laureys, S.; Celesia, G.G.; Cohadon, F.; Lavrijsen, J.; Leon-Carrion, J.; Sannita, W.G.; Sazbon, L.; Schmutzhard, E.; von Wild, K.R.; Zeman, A., et al. Unresponsive wakefulness syndrome: a new name for the vegetative state or apallic syndrome. Bmc Medicine 2010, 8, doi:10.1186/1741-7015-8-68.

3. Giacino, J.T.; Malone, R. The vegetative and minimally conscious states. Handb Clin Neurol 2008, 90, 99–111, doi:10.1016/S0072-9752(07)01706-X.

4. Bruno, M.A.; Vanhaudenhuyse, A.; Thibaut, A.; Moonen, G.; Laureys, S. From unresponsive wakefulness to minimally conscious PLUS and functional locked-in syndromes: recent advances in our understanding of disorders of consciousness. J Neurol 2011, 258, 1373–1384, doi:10.1007/s00415-011-6114-x.

5. Teasdale, G.; Jennett, B. Assessment of coma and impaired consciousness. A practical scale. Lancet 1974, 2, 81–84, doi:10.1016/s0140-6736(74)91639-0.

6. Kalmar, K.; Giacino, J.T. The JFK Coma Recovery Scale - revised. Neuropsychological Rehabilitation 2005, 15, 454–460, doi:Doi 10.1080/09602010443000425.

7. Blume, C.; Angerer, M.; Raml, M.; del Giudice, R.; Santhi, N.; Pichler, G.; Kunz, A.B.; Scarpatetti, M.; Trinka, E.; Schabus, M. Healthier rhythm, healthier brain? Integrity of circadian melatonin and temperature rhythms relates to the clinical state of brain-injured patients. European Journal of Neurology 2019, 26, 1051–1059, doi:10.1111/ene.13935.

8. Andrews, K.; Murphy, L.; Munday, R.; Littlewood, C. Misdiagnosis of the vegetative state: Retrospective study in a rehabilitation unit. Brit Med J 1996, 313, 13–16, doi:DOI 10.1136/bmj.313.7048.13.

9. Demertzi, A.; Antonopoulos, G.; Heine, L.; Voss, H.U.; Crone, J.S.; de Los Angeles, C.; Bahri, M.A.; Di Perri, C.; Vanhaudenhuyse, A.; Charland-Verville, V., et al. Intrinsic functional connectivity differentiates minimally conscious from unresponsive patients. Brain 2015, 138, 2619–2631, doi:10.1093/brain/awv169.

10. Stender, J.; Gosseries, O.; Bruno, M.A.; Charland-Verville, V.; Vanhaudenhuyse, A.; Demertzi, A.; Chatelle, C.; Thonnard, M.; Thibaut, A.; Heine, L., et al. Diagnostic precision of PET imaging and functional MRI in disorders of consciousness: a clinical validation study. Lancet 2014, 384, 514–522, doi:10.1016/S0140-6736(14)60042-8.

11. Monti, M.M.; Vanhaudenhuyse, A.; Coleman, M.R.; Boly, M.; Pickard, J.D.; Tshibanda, L.; Owen, A.M.; Laureys, S. Willful Modulation of Brain Activity in Disorders of Consciousness. New Engl J Med 2010, 362, 579–589, doi:10.1056/NEJMoa0905370.

12. Riganello, F.; Larroque, S.K.; Bahri, M.A.; Heine, L.; Martial, C.; Carriere, M.; Charland-Verville, V.; Aubinet, C.; Vanhaudenhuyse, A.; Chatelle, C., et al. A Heartbeat Away From Consciousness: Heart Rate Variability Entropy Can Discriminate Disorders of Consciousness and Is Correlated With Resting-State fMRI Brain Connectivity of the Central Autonomic Network. Front Neurol 2018, 9, 769, doi:10.3389/fneur.2018.00769.

13. Shaffer, F.; Ginsberg, J.P. An Overview of Heart Rate Variability Metrics and Norms. Front Public Health 2017, 5, 258, doi:10.3389/fpubh.2017.00258.

14. Thayer, J.F.; Hansen, A.L.; Saus-Rose, E.; Johnsen, B.H. Heart Rate Variability, Prefrontal Neural Function, and Cognitive Performance: The Neurovisceral Integration Perspective on Self-regulation, Adaptation, and Health. Ann Behav Med 2009, 37, 141–153, doi:10.1007/s12160-009-9101-z.

15. Gujjar, A.R.; Sathyaprabha, T.N.; Nagaraja, D.; Thennarasu, K.; Pradhan, N. Heart rate variability and outcome in acute severe stroke: role of power spectral analysis. Neurocrit Care 2004, 1, 347–353, doi:10.1385/NCC:1:3:347.

16. Rapenne, T.; Moreau, D.; Lenfant, F.; Vernet, M.; Boggio, V.; Cottin, Y.; Freysz, M. Could heart rate variability predict outcome in patients with severe head injury? A pilot study. J Neurosurg Anesthesiol 2001, 13, 260–268, doi:10.1097/00008506-200107000-00016.

17. Sara, M.; Sebastiano, F.; Sacco, S.; Pistoia, F.; Onorati, P.; Albertini, G.; Carolei, A. Heart rate non linear dynamics in patients with persistent vegetative state: a preliminary report. Brain Inj 2008, 22, 33–37, doi:10.1080/02699050701810670.

18. Garbarino, S.; Lanteri, P.; Feeling, N.R.; Jarczok, M.N.; Quintana, D.S.; Koenig, J.; Sannita, W.G. Circadian Rhythms, Sleep, and the Autonomic Nervous System. J Psychophysiol 2020, 34, 1–9, doi:10.1027/0269-8803/a000236.

19. Bilan, A.; Witczak, A.; Palusinski, R.; Myslinski, W.; Hanzlik, J. Circadian rhythm of spectral indices of heart rate variability in healthy subjects. J Electrocardiol 2005, 38, 239–243, doi:10.1016/j.jelectrocard.2005.01.012.

20. Boudreau, P.; Yeh, W.H.; Dumont, G.A.; Boivin, D.B. A circadian rhythm in heart rate variability contributes to the increased cardiac sympathovagal response to awakening in the morning. Chronobiol Int 2012, 29, 757–768, doi:10.3109/07420528.2012.674592.

21. Blume, C.; Lechinger, J.; Santhi, N.; del Giudice, R.; Gnjezda, M.T.; Pichler, G.; Scarpatetti, M.; Donis, J.; Michitsch, G.; Schabus, M. Significance of circadian rhythms in severely brain-injured patients A clue to consciousness? Neurology 2017, 88, 1933–1941, doi:10.1212/Wnl.0000000000003942.

22. Angerer, M.; Schabus, M.; Raml, M.; Pichler, G.; Kunz, A.B.; Scarpatetti, M.; Trinka, E.; Blume, C. Actigraphy in brain-injured patients - A valid measurement for assessing circadian rhythms? Bmc Medicine 2020, 18, doi:10.1186/s12916-020-01569-y.

23. Czeisler, C.A.; Gooley, J.J. Sleep and circadian rhythms in humans. Cold Spring Harb Symp Quant Biol 2007, 72, 579–597, doi:10.1101/sqb.2007.72.064.

24. Voss, A.; Schulz, S.; Schroeder, R.; Baumert, M.; Caminal, P. Methods derived from nonlinear dynamics for analysing heart rate variability. Philos Trans A Math Phys Eng Sci 2009, 367, 277–296, doi:10.1098/rsta.2008.0232.

25. Sheng, H.; Chen, Y.Q.; Qiu, T. On the robustness of Hurst estimators. Iet Signal Process 2011, 5, 209–225, doi:10.1049/iet-spr.2009.0241.

26. Wislowska, M.; Del Giudice, R.; Lechinger, J.; Wielek, T.; Heib, D.P.J.; Pitiot, A.; Pichler, G.; Michitsch, G.; Donis, J.; Schabus, M. Night and day variations of sleep in patients with disorders of consciousness. Sci Rep 2017, 7, 266, doi:10.1038/s41598-017-00323-4.

27. Angerer, M.; Pichler, G.; Angerer, B.; Scarpatetti, M.; Schabus, M.; Blume, C. From Dawn to Dusk – Mimicking Natural Daylight Exposure Improves Circadian Rhythm Entrainment in Patients with Severe Brain Injury. Under review, preprint: https://osf.io/vk2d6/.

28. Blechert, J.; Peyk, P.; Liedlgruber, M.; Wilhelm, F.H. ANSLAB: Integrated multichannel peripheral biosignal processing in psychophysiological science. Behav Res Methods 2016, 48, 1528–1545, doi:10.3758/s13428-015-0665-1.

29. Berntson, G.G.; Stowell, J.R. ECG artifacts and heart period variability: Don’t miss a beat! Psychophysiology 1998, 35, 127–132, doi:Doi 10.1111/1469-8986.3510127.

30. Wilhelm, F.H.; Grossman, P.; Roth, W.T. Assessment of heart rate variability during alterations in stress: complex demodulation vs. spectral analysis. Biomed Sci Instrum 2005, 41, 346–351.

31. Hayano, J.; Taylor, J.A.; Yamada, A.; Mukai, S.; Hori, R.; Asakawa, T.; Yokoyama, K.; Watanabe, Y.; Takata, K.; Fujinami, T. Continuous Assessment of Hemodynamic Control by Complex Demodulation of Cardiovascular Variability. American Journal of Physiology 1993, 264, H1229–H1238, doi:DOI 10.1152/ajpheart.1993.264.4.H1229.

32. R Core Team R: A language and environment for statistical computing., Vienna, Austria, 2015.

33. Asehnoune, K.; Roquilly, A.; Cinotti, R. Respiratory Management in Patients with Severe Brain Injury. Crit Care 2018, 22, doi:10.1186/s13054-018-1994-0.

34. Grossman, P.; Karemaker, J.; Wieling, W. Prediction of Tonic Parasympathetic Cardiac Control Using Respiratory Sinus Arrhythmia - the Need for Respiratory Control. Psychophysiology 1991, 28, 201–216, doi:DOI 10.1111/j.1469-8986.1991.tb00412.x.

35. Wasserstein, R.L.; Schirm, A.L.; Lazar, N.A. Moving to a World Beyond “p < 0.05”. Am Stat 2019, 73, 1–19, doi:10.1080/00031305.2019.1583913.

36. Bürkner, P.C. brms: An R Package for Bayesian Multilevel Models Using Stan. J Stat Softw 2017, 80, 1–28, doi:10.18637/jss.v080.i01.

37. Carpenter, B.; Gelman, A.; Hoffman, M.D.; Lee, D.; Goodrich, B.; Betancourt, M.; Riddell, A.; Guo, J.Q.; Li, P.; Riddell, A. Stan: A Probabilistic Programming Language. J Stat Softw 2017, 76, 1–29, doi:10.18637/jss.v076.i01.

38. Friedrich, S.; Konietschke, F.; Pauly, M. Resampling-Based Analysis of Multivariate Data and Repeated Measures Designs with the R Package MANOVA.RM. R J 2019, 11, 380–400.

39. Benjamini, Y.; Hochberg, Y. Controlling the False Discovery Rate - a Practical and Powerful Approach to Multiple Testing. J R Stat Soc B 1995, 57, 289–300, doi:DOI 10.1111/j.2517-6161.1995.tb02031.x.

40. Cugini, P.; Curione, M.; Cammarota, C.; Bernardini, F.; Cipriani, D.; Rosa, R.D.; Francia, P.; Laurentis, T.D.; Marco, E.D.; Napoli, A., et al. Is a Reduced Entropy in Heart Rate Variability an Early Finding of Silent Cardiac Neurovegetative Dysautonomia in Type 2 Diabetes Mellitus. Journal of clinical and basic cardiology 2001, 4, 289–294.

41. Byun, S.; Kim, A.Y.; Jang, E.H.; Kim, S.; Choi, K.W.; Yu, H.Y.; Jeon, H.J. Entropy analysis of heart rate variability and its application to recognize major depressive disorder: A pilot study. Technol Health Care 2019, 27, S407–S424, doi:10.3233/Thc-199037.

42. Lechinger, J.; Bothe, K.; Pichler, G.; Michitsch, G.; Donis, J.; Klimesch, W.; Schabus, M. CRS-R score in disorders of consciousness is strongly related to spectral EEG at rest. J Neurol 2013, 260, 2348–2356, doi:10.1007/s00415-013-6982-3.

43. Batchinsky, A.L.; Cancio, L.C.; Salinas, J.; Kuusela, T.; Cooke, W.H.; Wang, J.J.; Boehme, M.; Convertino, V.A.; Holcomb, J.B. Prehospital loss of R-to-R interval complexity is associated with mortality in trauma patients. J Trauma 2007, 63, 512–518, doi:10.1097/TA.0b013e318142d2f0.

44. Gao, L.; Smielewski, P.; Czosnyka, M.; Ercole, A. Cerebrovascular Signal Complexity Six Hours after Intensive Care Unit Admission Correlates with Outcome after Severe Traumatic Brain Injury. J Neurotraum 2016, 33, 2011–2018, doi:10.1089/neu.2015.4228.

45. Bagnato, S.; Boccagni, C.; Sant’Angelo, A.; Fingelkurts, A.A.; Fingelkurts, A.A.; Galardi, G. Longitudinal Assessment of Clinical Signs of Recovery in Patients with Unresponsive Wakefulness Syndrome after Traumatic or Nontraumatic Brain Injury. J Neurotraum 2017, 34, 535–539, doi:10.1089/neu.2016.4418.

46. Luaute, J.; Maucort-Boulch, D.; Tell, L.; Quelard, F.; Sarraf, T.; Iwaz, J.; Boisson, D.; Fischer, C. Long-term outcomes of chronic minimally conscious and vegetative states. Neurology 2010, 75, 246–252, doi:DOI 10.1212/WNL.0b013e3181e8e8df.

47. Katz, D.I.; Polyak, M.; Coughlan, D.; Nichols, M.; Roche, A. Natural history of recovery from brain injury after prolonged disorders of consciousness: outcome of patients admitted to inpatient rehabilitation with 1-4 year follow-up. Prog Brain Res 2009, 177, 73–88, doi:10.1016/S0079-6123(09)17707-5.

48. Echeverria, J.C.; Woolfson, M.S.; Crowe, J.A.; Hayes-Gill, B.R.; Croaker, G.D.; Vyas, H. Interpretation of heart rate variability via detrended fluctuation analysis and alphabeta filter. Chaos 2003, 13, 467–475, doi:10.1063/1.1562051.

49. Goldberger, A.L. Is the Normal Heartbeat Chaotic or Homeostatic. News Physiol Sci 1991, 6, 87–91, doi:DOI 10.1152/physiologyonline.1991.6.2.87.

