## Supplementary Material for "Does the Heart Fall Asleep? – Diurnal Variations of Heart Rate Variability in Patients with Disorders of Consciousness"

### Missing Data

**Table S1.** Amount of missing data from each subject separately for time (i.e., forenoon [8am-2pm], afternoon [2pm-8pm], night [11pm-5am]).

| | Forenoon | | Afternoon | | Night | |
|---|---|---|---|---|---|---|
| **Patient ID** | **Frequency** | **Percentage** | **Frequency** | **Percentage** | **Frequency** | **Percentage** |
| P1 | 0 | 0 | 0 | 0 | 0 | 0 |
| P2 | 23 | 6.39 | 0 | 0 | 0 | 0 |
| P3 | 6 | 1.67 | 0 | 0 | 0 | 0 |
| P4 | 7 | 1.94 | 0 | 0 | 0 | 0 |
| P5 | 0 | 0 | 0 | 0 | 0 | 0 |
| P6 | 0 | 0 | 21 | 5.83 | 0 | 0 |
| P7 | 0 | 0 | 10 | 2.78 | 0 | 0 |
| P8 | 0 | 0 | 0 | 0 | 26 | 7.22 |
| P9 | 0 | 0 | 0 | 0 | 0 | 0 |
| P10 | 1 | 0.28 | 0 | 0 | 0 | 0 |
| P11 | 2 | 0.56 | 0 | 0 | 0 | 0 |
| P12 | 0 | 0 | 0 | 0 | 0 | 0 |
| P13 | 0 | 0 | 0 | 0 | 0 | 0 |
| P14 | 0 | 0 | 0 | 0 | 0 | 0 |
| P15 | 0 | 0 | 0 | 0 | 0 | 0 |
| P16 | 0 | 0 | 0 | 0 | 0 | 0 |
| P17 | 0 | 0 | 0 | 0 | 0 | 0 |
| P18 | 0 | 0 | 0 | 0 | 0 | 0 |
| P19 | 0 | 0 | 0 | 0 | 0 | 0 |
| P20 | 0 | 0 | 0 | 0 | 0 | 0 |
| P21 | 0 | 0 | 0 | 0 | 0 | 0 |
| P22 | 0 | 0 | 0 | 0 | 0 | 0 |
| P23 | 0 | 0 | 0 | 0 | 0 | 0 |
| P24 | 0 | 0 | 0 | 0 | 0 | 0 |
| P25 | 0 | 0 | 0 | 0 | 0 | 0 |
| P26 | 0 | 0 | 0 | 0 | 0 | 0 |

Frequency refers to the amount of one-minute segments that are missing. Percentage describes the amount of missing one-minute segments in its relation to the total file length of six hours (i.e., 360 one-minute segments). Missing data is highlighted in grey.

### Normality Test Results

**Table S2.** Shapiro-Wilk tests ($p$-values) for normality for each variable separately for time (i.e., forenoon [8am-2pm], afternoon [2pm-8pm], night [11pm-5am]).

| **Variable** | **Forenoon** | **Afternoon** | **Night** |
|---|---|---|---|
| Interbeat interval | .436 | .551 | .729 |
| Heart rate | .054 | .061 | .025 |
| RMSSD | .013 | .003 | .194 |
| Very low frequency | .012 | .075 | .281 |
| Low frequency | .005 | .017 | .497 |
| High frequency | .001 | .008 | .004 |
| Approximate entropy | .907 | .162 | .193 |

| | | | | |
|---|---|---|---|---|
| DfaAlpha | | .067 | .002 | .352 |
| Hurst exponent | | .162 | .056 | .061 |
| Sample entropy 1 | | .001 | < .001 | < .001 |
| Sample entropy 2 | | .212 | .703 | .469 |
| Sample entropy 3 | | .054 | .693 | .603 |
| Sample entropy 4 | | .092 | .687 | .705 |
| Sample entropy 5 | | .079 | .652 | .916 |
| EEG permutation entropy | | | .219 | .681 |
| CRS-R sum score | < .001 | | | |
| CRS-R auditory subscale score | < .001 | | | |
| CRS-R visual subscale score | < .001 | | | |
| CRS-R motor subscale score | < .001 | | | |
| CRS-R oroverbal subscale score | < .001 | | | |
| CRS-R communication subscale score | < .001 | | | |
| CRS-R arousal subscale score | < .001 | | | |
| Age | < .001 | | | |
| Time since injury | < .001 | | | |

*P*-values for EEG permutation entropy refer to day (i.e., 8am-8pm) and night (i.e., 11pm-5am). *P*-values for CRS-R scores, age and time since injury refer to all time conditions (i.e., forenoon, afternoon, and night). Significant *p*-values (i.e. $p < .05$), which indicate that the data is not normal distributed, are marked in grey. Abbreviations: RMSSD = root mean square of successive differences between adjacent heartbeats, DfaAlpha = detrended fluctuation analysis scaling exponent.

### Results

#### *Heart Rate*

Analyses of the heart rate (HR) of 26 patients revealed a trend towards a main effect for *time* ($F_{WTS}(2)=7.28$, $p=.053$) and a significant effect for *diagnosis* ($F_{WTS}(1)=5.1$, $p=.039$), but no significant *time* × *diagnosis* interaction ($F_{WTS}(2)=1.42$, $p=.515$). Specifically, patients showed a lower HR during night as compared to forenoon ($F_{WTS}(1)=8.26$, $p=.021$) and afternoon ($F_{WTS}(1)=6.64$, $p=.024$). No differences could be observed in the patients' HR between fore- and afternoon ($F_{WTS}(1)=0.002$, $p=.967$; cf. *Figure S1a*). Further, patients with UWS showed a lower HR as compared to patients with (E)MCS ($F_{WTS}(1)=5.1$, $p=.039$; cf. *Figure S1b*). This effect was also visible when correlating HR and CRS-R sum scores of 25 patients, showing that lower CRS-R sum scores were associated with lower HR during forenoon ($r_{\tau}(23)=0.34$, $p=.02$; cf. *Figure S2a*), afternoon ($r_{\tau}(23)=0.38$, $p=.009$; cf. *Figure S2b*) and night ($r_{\tau}(23)=0.28$, $p=.056$; $b= 1.41$, 95% CI = [0.10, 2.72]; cf. *Figure S2c*).

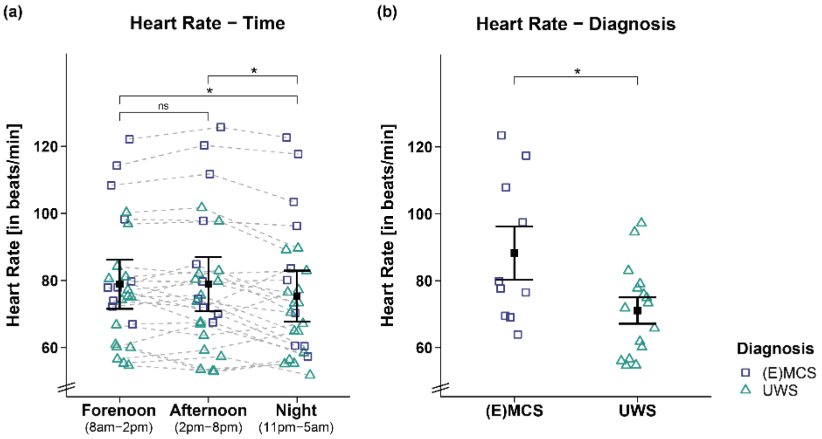


**Figure S1.** Heart rate (HR) separately for time and diagnosis contrasts. **(a)** While patients' HR was lower during the night as compared to the day (i.e., forenoon, afternoon), it did not differ between fore- and afternoon. **(b)** Patients with UWS showed a lower HR than patients with (E)MCS. Error bars represent the mean and 95% confidence interval. *$p$ < .05, ns = not significant. Abbreviations: (E)MCS = (emergence from) minimally conscious state, UWS = unresponsive wakefulness syndrome, beats/min = heartbeats per minute.

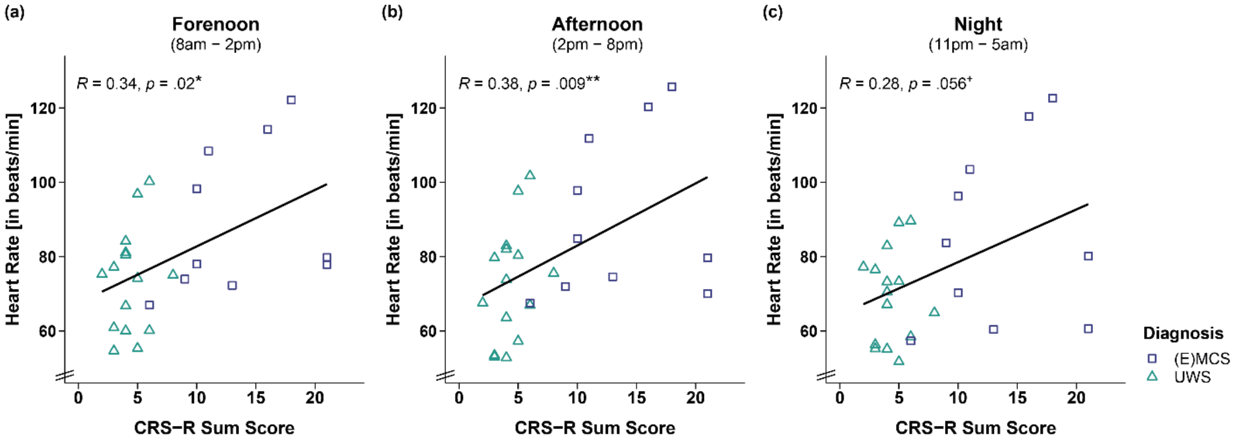


**Figure S2. Correlation between heart rate (HR) and CRS-R sum score separately for time.** A higher HR was associated with a higher CRS-R sum score throughout the **(a, b)** day (i.e., forenoon, afternoon) and **(c)** night. **$p$ < .01, *$p$ < .05, $^{+}p \leq .1$. Abbreviations: (E)MCS = (emergence from) minimally conscious state, UWS = unresponsive wakefulness syndrome, beats/min = heartbeats per minute.

#### *Etiology*

**Table S3.** Wald-type statistic (WTS) of the different HRV parameters separately for the main effects 'time' (i.e., forenoon [8am-2pm], afternoon [2pm-8pm], night [11pm-5am]) and 'etiology' (i.e., traumatic vs. non-traumatic brain injury), and the 'time × etiology' interaction ($N$=26).

| Variable | Time | Etiology | Time × Etiology |
|---|---|---|---|
| Interbeat interval | $F_{WTS}(2)=7.37$, $p$=.050 | $F_{WTS}(1)=0.08$, $p$=.778 | $F_{WTS}(2)=5.58$, $p$=.088 |
| Heart rate | $F_{WTS}(2)=7.82$, $p$=.041 | $F_{WTS}(1)=0.00$, $p$=.949 | $F_{WTS}(2)=3.33$, $p$=.220 |
| RMSSD | $F_{WTS}(2)=0.50$, $p$=.789 | $F_{WTS}(1)=0.00$, $p$=.961 | $F_{WTS}(2)=1.51$, $p$=.502 |
| Very low frequency | $F_{WTS}(2)=7.47$, $p$=.046 | $F_{WTS}(1)=0.22$, $p$=.648 | $F_{WTS}(2)=2.19$, $p$=.368 |
| Low frequency | $F_{WTS}(2)=6.59$, $p$=.065 | $F_{WTS}(1)=0.02$, $p$=.893 | $F_{WTS}(2)=2.90$, $p$=.268 |
| High frequency | $F_{WTS}(2)=4.08$, $p$=.178 | $F_{WTS}(1)=0.24$, $p$=.619 | $F_{WTS}(2)=2.16$, $p$=.382 |
| Approximate entropy | $F_{WTS}(2)=20.74$, $p$=.001 | $F_{WTS}(1)=0.30$, $p$=.575 | $F_{WTS}(2)=2.90$, $p$=.277 |
| DfaAlpha | $F_{WTS}(2)=0.44$, $p$=.816 | $F_{WTS}(1)=4.59$, $p$=.043 | $F_{WTS}(2)=1.29$, $p$=.552 |
| Hurst exponent | $F_{WTS}(2)=2.36$, $p$=.335 | $F_{WTS}(1)=0.09$, $p$=.772 | $F_{WTS}(2)=2.01$, $p$=.400 |
| Sample entropy 1 | $F_{WTS}(2)=0.41$, $p$=.830 | $F_{WTS}(1)=4.36$, $p$=.049 | $F_{WTS}(2)=1.22$, $p$=.579 |
| Sample entropy 2 | $F_{WTS}(2)=3.79$, $p$=.193 | $F_{WTS}(1)=0.51$, $p$=.477 | $F_{WTS}(2)=2.04$, $p$=.396 |
| Sample entropy 3 | $F_{WTS}(2)=3.4$, $p$=.226 | $F_{WTS}(1)=1.03$, $p$=.316 | $F_{WTS}(2)=1.34$, $p$=.543 |
| Sample entropy 4 | $F_{WTS}(2)=3.26$, $p$=.239 | $F_{WTS}(1)=1.07$, $p$=.304 | $F_{WTS}(2)=0.68$, $p$=.734 |
| Sample entropy 5 | $F_{WTS}(2)=2.43$, $p$=.338 | $F_{WTS}(1)=1.09$, $p$=.300 | $F_{WTS}(2)=0.78$, $p$=.701 |

Significant $p$-values (i.e. $p$ < .05) are marked in grey. Abbreviations: RMSSD = root mean square of successive differences between adjacent heartbeats, DfaAlpha = detrended fluctuation analysis scaling exponent.

#### *Sex*

**Table S4.** Wald-type statistic (WTS) of the different HRV parameters separately for the main effects 'time' (i.e., forenoon [8am-2pm], afternoon [2pm-8pm], night [11pm-5am]), 'sex' (i.e., male vs. female), and the "time × sex" interaction ($N$=26).

| Variable | Time | Sex | Time × Sex |
|---|---|---|---|
| Interbeat interval | $F_{WTS}(2)=4.66$, $p$=.156 | $F_{WTS}(1)=2.36$, $p$=.143 | $F_{WTS}(2)=3.89$, $p$=.205 |

| | | | |
|---|---|---|---|
| Heart rate | $F_{WTS}(2)=6.06$, $p=.099$ | $F_{WTS}(1)=1.96$, $p=.176$ | $F_{WTS}(2)=3.82$, $p=.217$ |
| RMSSD | $F_{WTS}(2)=0.18$, $p=.920$ | $F_{WTS}(1)=0.44$, $p=.514$ | $F_{WTS}(2)=1.41$, $p=.547$ |
| Very low frequency | $F_{WTS}(2)=12.21$, $p=.014$ | $F_{WTS}(1)=0.09$, $p=.773$ | $F_{WTS}(2)=0.11$, $p=.950$ |
| Low frequency | $F_{WTS}(2)=12.08$, $p=.015$ | $F_{WTS}(1)=0.53$, $p=.478$ | $F_{WTS}(2)=0.05$, $p=.977$ |
| High frequency | $F_{WTS}(2)=1.1$, $p=.622$ | $F_{WTS}(1)=1.45$, $p=.242$ | $F_{WTS}(2)=1.82$, $p=.310$ |
| Approximate entropy | $F_{WTS}(2)=22.29$, $p=.002$ | $F_{WTS}(1)=0.26$, $p=.607$ | $F_{WTS}(2)=2.27$, $p=.383$ |
| DfaAlpha | $F_{WTS}(2)=0.05$, $p=.975$ | $F_{WTS}(1)=3.92$, $p=.062$ | $F_{WTS}(2)=4.89$, $p=.135$ |
| Hurst exponent | $F_{WTS}(2)=1.59$, $p=.495$ | $F_{WTS}(1)=0.45$, $p=.507$ | $F_{WTS}(2)=8.47$, $p=.044$ |
| Sample entropy 1 | $F_{WTS}(2)=0.3$, $p=.883$ | $F_{WTS}(1)=1.45$, $p=.253$ | $F_{WTS}(2)=2.78$, $p=.325$ |
| Sample entropy 2 | $F_{WTS}(2)=4.51$, $p=.159$ | $F_{WTS}(1)=0.00$, $p=.985$ | $F_{WTS}(2)=2.17$, $p=.394$ |
| Sample entropy 3 | $F_{WTS}(2)=3.57$, $p=.229$ | $F_{WTS}(1)=0.04$, $p=.858$ | $F_{WTS}(2)=1.89$, $p=.443$ |
| Sample entropy 4 | $F_{WTS}(2)=3.19$, $p=.268$ | $F_{WTS}(1)=0.05$, $p=.824$ | $F_{WTS}(2)=1.41$, $p=.537$ |
| Sample entropy 5 | $F_{WTS}(2)=2.01$, $p=.428$ | $F_{WTS}(1)=0.08$, $p=.792$ | $F_{WTS}(2)=1.39$, $p=.541$ |

Abbreviations: RMSSD = root mean square of successive differences between adjacent heartbeats, DfaAlpha = detrended fluctuation analysis scaling exponent.

*Entropy Parameters*

**Table S5.** Wald-type statistic (WTS) of the different entropy parameters separately for the main effects 'time' (i.e., forenoon [8am-2pm], afternoon [2pm-8pm], night [11pm-5am]) and 'diagnosis' (i.e., E/MCS vs. UWS), and the 'time × diagnosis' interaction ($N$=26).

| Variable | Time | Diagnosis | Time × Diagnosis |
|---|---|---|---|
| DfaAlpha | $F_{WTS}(2)=0.48$, $p=.807$ | $F_{WTS}(1)=1.53$, $p=.228$ | $F_{WTS}(2)=0.71$, $p=.721$ |
| Hurst exponent | $F_{WTS}(2)=3.05$, $p=.257$ | $F_{WTS}(1)=0.22$, $p=.646$ | $F_{WTS}(2)=1.08$, $p=.604$ |
| Sample entropy 1 | $F_{WTS}(2)=0.40$, $p=.841$ | $F_{WTS}(1)=0.44$, $p=.518$ | $F_{WTS}(2)=0.24$, $p=.901$ |
| Sample entropy 2 | $F_{WTS}(2)=4.27$, $p=.161$ | $F_{WTS}(1)=0.07$, $p=.791$ | $F_{WTS}(2)=0.05$, $p=.980$ |
| Sample entropy 3 | $F_{WTS}(2)=3.97$, $p=.178$ | $F_{WTS}(1)=0.01$, $p=.941$ | $F_{WTS}(2)=0.06$, $p=.972$ |
| Sample entropy 4 | $F_{WTS}(2)=3.71$, $p=.197$ | $F_{WTS}(1)=0.12$, $p=.737$ | $F_{WTS}(2)=0.09$, $p=.958$ |
| Sample entropy 5 | $F_{WTS}(2)=2.93$, $p=.271$ | $F_{WTS}(1)=0.21$, $p=.650$ | $F_{WTS}(2)=0.01$, $p=.996$ |

Abbreviation: DfaAlpha = detrended fluctuation analysis scaling exponent.

*EEG Entropy*

Analyses of the EEG permutation entropy (PE) of 14 patients revealed a significant main effect for *time* ($F_{WTS}(2)=9.79$, $p=.011$) with patients showing higher PE during the day (i.e., 8am-8pm) as compared to the night (i.e., 11pm-5am), and a significant *time × diagnosis* interaction ($F_{WTS}(2)=5.39$, $p=.041$), but no significant main effect for *diagnosis* ($F_{WTS}(1)=0.12$, $p=.733$). Specifically, while patients with (E)MCS showed a lower PE during the night than during the day ($F_{WTS}(1)=21.87$, $p=.002$), no such day-night difference was observed in patients with UWS ($F_{WTS}(1)=0.25$, $p=.631$). This has already been shown in [25]. We ran the analyses again as we only use a subsample of the dataset here.
